# The clinical, genomic, and microbiological profile of invasive multi-drug resistant Escherichia coli in a major teaching hospital in the United Kingdom

**DOI:** 10.1101/2023.06.01.23290836

**Authors:** William L. Hamilton, Suny Coscione, Mailis Maes, Ben Warne, Lindsay J. Pike, Fahad A. Khokhar, Beth Blane, Nicholas M. Brown, Theodore Gouliouris, Gordon Dougan, M. Estée Török, Stephen Baker

## Abstract

*Escherichia coli* is a ubiquitous component of the human gut microbiome, but is also a common pathogen, causing around 40,000 bloodstream infections (BSI) in the UK annually. The number of *E. coli* BSI has increased over the last decade in the UK, and emerging antimicrobial resistance (AMR) profiles threaten treatment options. Here, we combined clinical, epidemiological, and whole genome sequencing data with high content imaging to characterise over 300 *E. coli* isolates associated with BSI in a large UK teaching hospital. Overall, only a limited number of sequence types (ST) were responsible for the majority of organisms causing invasive disease. The most abundant (20% of all isolates) was ST131, of which around 90% comprised the pandemic O25:H4 group. ST131-O25:H4 isolates were frequently multi-drug resistant (MDR), with a high prevalence of extended spectrum β-lactamases (ESBL) and fluoroquinolone resistance. There was no association between AMR phenotypes and the source of *E. coli* bacteraemia or whether the infection was healthcare-associated. Several clusters of ST131 were genetically similar, potentially suggesting a shared transmission network. However, there was no clear epidemiological associations between these cases, and they included organisms from both healthcare-associated and non-healthcare-associated origins. The majority of ST131 genetic clusters exhibited strong binding with an anti-O25b antibody, raising the possibility of developing rapid diagnostics targeting this pathogen. In summary, our data suggest that a restricted set of MDR *E. coli* populations can be maintained and spread across both community and healthcare settings in this location, contributing disproportionately to invasive disease and AMR.

**Impact statement:** The pandemic ST131-O25:H4 lineage was a common cause of *E. coli* bloodstream infections in this location, and carried multiple AMR mechanisms including ESBL and fluoroquinolone resistance. The conserved antigenicity of ST131-O25:H4 raises the possibility of developing targeted immune therapeutics and rapid diagnostics for this common and frequently multi-drug resistant pathogen.

## Introduction

*Escherichia coli* (*E. coli*) is a ubiquitous commensal of the human gut that can also cause a spectrum of invasive disease [1]. Globally, out of an estimated 13.7 million infection-related deaths in 2019, *E. coli* caused the second highest number of deaths among 33 investigated bacterial pathogens, at 950,000 deaths (after Staphylococcus aureus with 1.1 million deaths) [2]. Bloodstream infections (BSI) caused by *E. coli* cause significant morbidity and mortality [2, 3], and the incidence is increasing year on year (Supplementary Figure 1) [4]. *E. coli* has become the commonest cause of BSI in the United Kingdom (UK), causing around 40,000 BSI in the UK annually [5]. BSI can trigger acute systemic inflammatory response syndromes that can cause severe end-organ damage and death. Therefore, the prompt recognition, diagnosis and initiation of effective treatment, including antimicrobials, is essential. Gram-negative bacteraemia is becoming increasingly recognised as a public health threat and interventions aimed at reducing its incidence, including infections caused by *E. coli*, are a strategic priority in the UK [6].

Antimicrobial resistance (AMR) limits the therapeutic options for many infections, including BSI caused by Gram negative bacteria such as *E. coli* [7]. As *E. coli* occupy the gut they are a common sink and source for AMR genes, which can catalyse resistance to a wide array of antimicobials [8–10]. There is evidence from a Danish cohort that *E. coli* plays a key role in driving the infant gut microbiome towards higher abundance of AMR-associated genes [11]. Understanding the production and proliferation of AMR among microbial communities is a key challenge in clinical microbiology. The relationships between *E. coli* genomic diversity, including the mobile genetic elements that can transport AMR genes, and their clinical and epidemiological characteristics, merit further investigation. These analyses require integration of genomics with clinical microbiology, which can be challenging in healthcare settings.

*E. coli* sequence type (ST)-131, serotype O25:H4, has spread internationally and is responsible for a significant burden of extended spectrum β-lactamase (ESBL) *E. coli* disease in the UK and globally, in both healthcare and community settings [12–19], in the food chain [20] and in animals [21]. This pandemic lineage possesses virulence mechanisms [22] and is associated with CTX-M-15 β-lactamase production and fluoroquinolone resistance. There is frequent movement of β-lactamase genes between plasmids and the chromosome, and even closely related ST131 isolates can harbour substantial variation in their complement of plasmid-mediated AMR genes [23]. The genetic architecture underlying AMR in *E. coli* ST131, and how this propagates between individuals and relates to both asymptomatic carriage and invasive disease, is poorly understood.

Antibacterial immune therapeutics and rapid diagnostic testing are promising tools against invasive bacterial disease, particularly in the context of spreading and intensifying AMR, and a shortage of novel antibacterial agents. Anti-O25b vaccines and diagnostics have been proposed to tackle ST131 [24–26]; however, their design and implementation relies on understanding the genetic variation present in bacterial target genes and populations. This emphasises the importance of linking genomics with clinical, microbiological and epidemiological data.

Here, we aimed to determine the clinical, genomic, and microbiological profile of invasive multi-drug resistant (MDR) *E. coli* in the UK, by studying patients with E.coli bloodstream infections in a major hospital in the East of England. All BSI *E. coli* isolates were whole-genome sequenced and underwent a detailed phylogenetic analysis to determine the genomic epidemiology of these infections in hospital and community settings. Lastly, exploiting a commercial monoclonal antibody (mAb) against the O25b antigen, we aimed to understand antigen conservation in ST131 isolates to add to data supporting the development of immune therapeutics and diagnostics for common AMR/MDR organisms associated with BSI.

## Methods

### Study setting and processing of clinical specimens

All blood culture isolates were collected from patients at Cambridge University Hospitals NHS Foundation Trust (CUH) in the East of England. Comprising Addenbrooke’s and the Rosie Hospitals, CUH has approximately 1,100 acute beds and provides secondary care services for Cambridge and the surrounding area, serving a population of approximately 580,000 people. It also provides tertiary care services (including infectious diseases, oncology, haematology, solid organ transplantation, neurosurgery, paediatrics and neonatology) for the East of England.

Blood culture sets (BacT/ALERT, bioMérieux) were obtained from patients as clinically indicated. All samples were processed in the UKHSA Clinical Microbiology and Public Health Laboratory (CMPHL), which operates from the CUH site. All routine microbiological techniques were performed according to UK Standards for Microbiology Investigations (UK SMIs) [27]. Species identification was determined using mass spectrometry (MALDI-TOF MS, Bruker Daltonics). Antimicrobial susceptibility testing was determined using the disc diffusion method, with breakpoints determined by the BSAC criteria in use at the time (this study pre-dates the laboratory change to EUCAST methodology). All blood culture isolates were prospectively stored in glycerol broth at −80°C.

*E. coli* isolates of interest were identified from a database of all culture results from CUH, exported from the CMPHL laboratory information system, and filtered based on sample type and species. This provided data on sample collection and result dates, patient identifiers and antimicrobial susceptibility results determined by the clinical laboratory. All available *E. coli* isolates from samples collected between 1st December 2016 and 31st December 2017 were included in the study.

### Genome sequencing

Whole-genome sequencing was performed on all available stored isolates, except for duplicate isolates defined as blood culture samples from the same patient occurring within 90 days of the first isolate collection date. Standard DNA extraction methods were used as described in Supplementary Methods. DNA libraries were prepared using the IHTP WGS Ultra II library kit and sequenced on an Illumina HiSeq platform (Illumina) using a standard protocol at the Wellcome Sanger Institute.

### Bioinformatics

#### QC filters

The following quality control (QC) filtering was applied to genome sequences: total genome length <6MB, contig count <150, Kraken organism 1 is *E. coli*, and % reads mapping to Kraken organism 1 >30% (Supplementary Figures 3-5). The resulting phylogenetic tree included a long branch with 4 outlier samples, which were removed for subsequent analyses to yield the final analysis set (Supplementary Figure 6).

#### Core genome alignment and producing phylogenetic tree

Core genome alignment, tree building, SNP distance matrix, AMR gene detection, plasmid detection, multilocus sequence typing and *in silico* OH serotyping were performed on the Wellcome Sanger Institute high performance computing (HPC) network. Downstream analyses and visualisations were produced using the R programming language with *tidyverse* packages. Further details can be found in Supplementary Methods. Briefly, core genome alignment was performed using *roary* [28], requiring genes to be shared across 99% of isolates (Supplementary Table 1). A maximum likelihood phylogenetic tree was generated from the core genome alignment using IQTREE [29], using a GTR+G substitution model [30] with ultrafast bootstrapping for branch support [31]. The resulting phylogenetic tree was initially visualised and manipulated using Microreact [32] and final figures presented in the paper were produced using *ggtree* [33]. A SNP distance matrix was produced using snp-dists [34].

#### AMR gene and plasmid detection

AMR genes were detected from the CARD database, downloaded from: https://card.mcmaster.ca/, version 3.1.1 [35]. ARIBA [36] was used to prepare the CARD database and analyse the samples. *plasmidfinder* was used to identify plasmids [37], also prepared using ARIBA.

#### Multilocus sequence typing (MLST) and OH serotyping

MLST typing and OH serotyping were performed using srst2 [38].

#### Clinical and epidemiological data linkage

Prior to pseudonymisation for sequencing, the patient Medical Record Number was used to link records to the CUH electronic health record system (Epic, Epic Systems) and to locally held infection control databases created for submission to the Public Health England (PHE, now UK Health Security Agency) mandatory surveillance programme for hospital acquired infections.

Demographic data including gender, age, main specialty, etc were extracted from Epic, whilst suspected infectious focus was obtained from the infection control database. Categorisation of infection (healthcare-onset, healthcare-associated or community-onset) was made by accessing individual patient records and making an assessment based on adapted criteria, described in Supplementary Methods. To assess epidemiological association between selected cases, bed movements during hospital admission were reviewed. Strong epidemiological linkage was defined as two patients having been on the same ward and bay at any stage during admission. Weak epidemiological linkage was defined as having been on the same ward or under the same clinical specialty up to a week apart, without close contact based on bed location.

#### Antibody binding and image analysis

The monoclonal antibody used for high-content antibody binding assay was KM467, an IgG1 antibody based on VH and VL sequences of 3E9-11 [39]. The synthesis of this antibody and methodology used for the high content imaging assay have been previously described [40]. The full protocol for antibody binding is described in Supplementary Methods. Briefly, bacterial colonies were picked, cultured overnight in LB broth, diluted, washed and incubated with the KM467 antibody in CellCarrier-96 Ultra plates (Perkin Elmer), followed by addition of anti-human IgG and DAPI. Plates were imaged on an Opera Phenix confocal microscope (PerkinElmer) using the Alexa Fluor 647 and DAPI channels with the 63x water immersion objective. 16 fields and 3 Z-stacks were imaged per well. The image analysis was carried out using PerkinElmer Harmony (v4.9). The final result output was categorical, based on whether there was antibody binding or no binding imaged on the plate.

#### Statistical analysis

Logistic regression to test for associations between variables and 30-day mortality were performed in R using a generalized linear model. The R code took the general form:

> model <-glm(mortality_30_days ∼.,family=binomial(link=’logit’), data=metadata)

> summary(model)

> anova(model, test=“Chisq”)

Where the ‘metadata’ file comprised columns of metadata such as age, sex, etc.

Clusters of ST131 were defined from the SNP difference matrix produced from the *snp-dists* package, using *scipy.cluster.hierarchy functions dendrogram, linkage*, and *fcluster* to identify clusters within a specified genetic distance from each-other, corresponding to the intended SNP difference.

#### Ethics

This study was conducted under the terms of ethical approval granted by the NHS Research Ethics Committee for the project “Whole genome sequencing of bacterial pathogens” (reference: 12/EE/0439).

## Results

### Study population and clinical outcome

A total of 451 *E. coli* BSI isolates at Cambridge University Hospitals NHS Foundation Trust were identified within the designated study dates (1st December 2016 to 31st December 2017). After sample de-duplication, there were 349 patients with *E. coli* BSI represented by at least one sample, and whole-genome sequencing was performed on at least one *E. coli* isolate originating from 338 patients (Figure 1). After genome quality control (QC) filtering, the final analysis group comprised 322 *E. coli* BSI isolates with high-quality whole genome sequence information and associated metadata.

**Figure 1.**
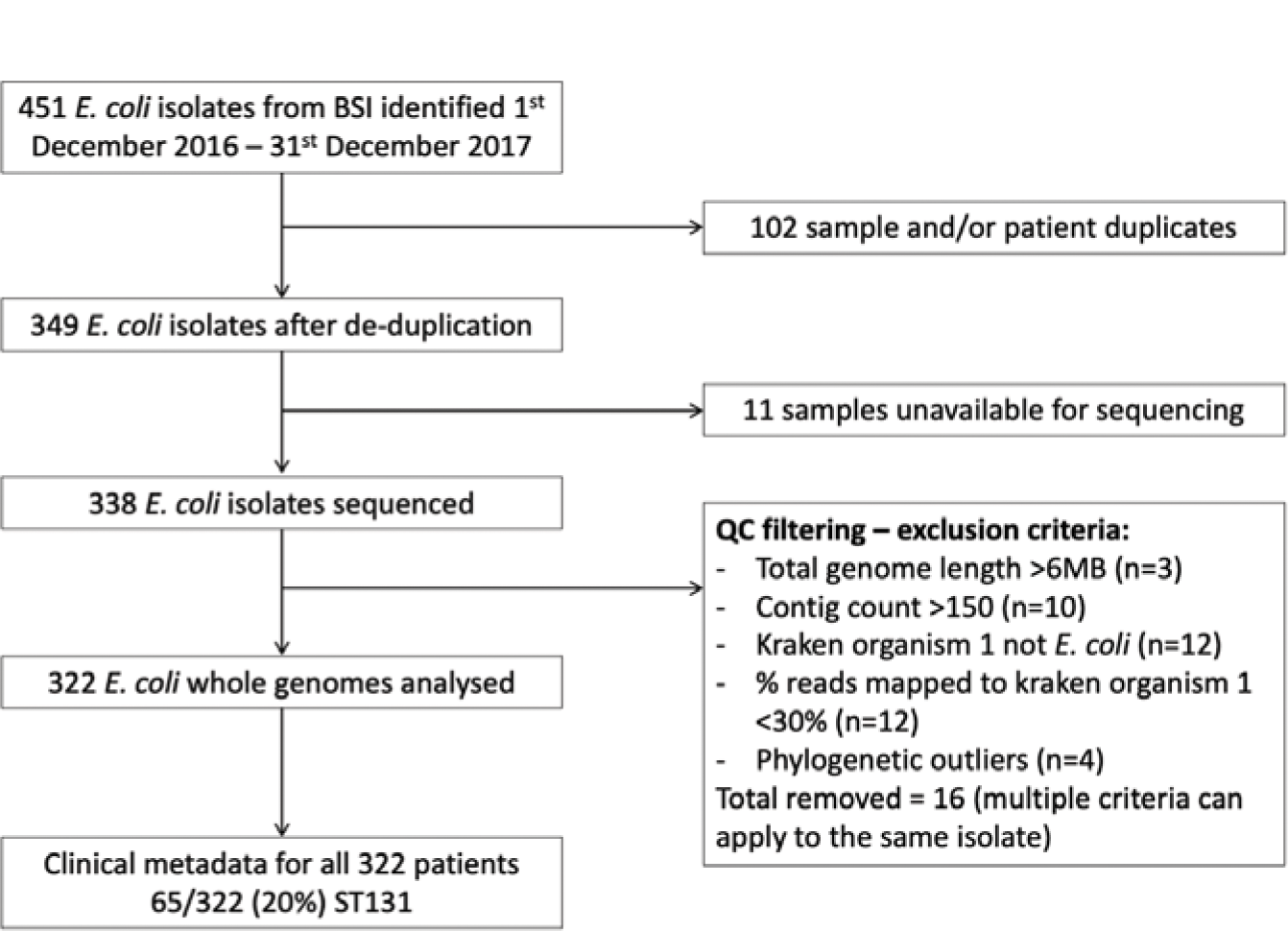
Study flow diagram. This shows the number of *E. coli* samples included in the study and in the final analysis. 451 *E. coli* isolates from 349 individuals were identified within the study period, of which 338 were sequenced and 322 passed sequencing QC filtering.

The epidemiological characteristics of the study population are shown in Table 1. Where patient address information was available (n=161/322, 50%), the majority of patients (118/161, 73%) lived within the local county (Cambridgeshire) (Supplementary Figure 2). The most commonly identified source of the bacteraemia (151/322; 47%) was a primary urinary tract infection (UTI). Overall, 128/322 (40%) of cases were categorised as being of ‘community-onset, healthcare-associated’; 105/322 (33%) were of ‘community-onset, non-healthcare-associated’, and 83/322 (26%) were of hospital-onset, healthcare-associated.

**Table 1.**
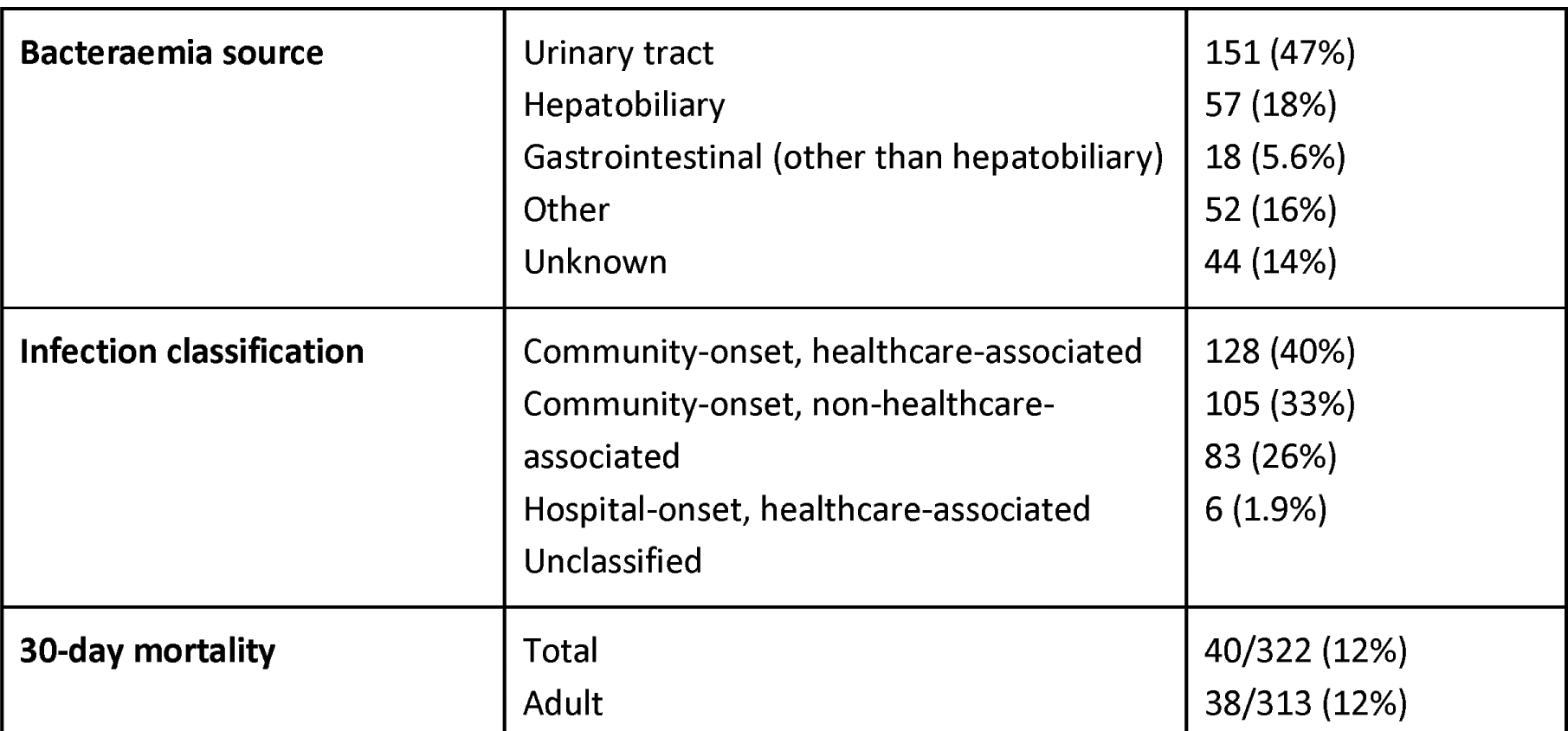
Study population epidemiological characteristics. Only patients associated with high quality *E. coli* genomes included in the study are presented. IQR = Interquartile Range.

In total, 40/322 (12%) patients died within 30 days of the onset of bacteraemia. The association between 30-day mortality in adult patients with age, sex, healthcare association, bacteraemia source, whether the *E. coli* had an ESBL or AmpC phenotype, and whether the organism was ST131, was investigated by logistic regression. Only age and bacteraemia source were significantly associated with 30-day mortality (*p*=0.049 and *p*=1.6×10, respectively). Overall, 18/111 (16%) adults aged >80 years old died within 30 days of BSI onset, compared with 20/202 (9.9%) in patients aged <80 years old (Supplementary Table 2). BSI with a urinary tract source was strongly associated with reduced odds of 30-day mortality: 5/149 (3.4%) adults with urinary tract source died within 30 days compared with 33/164 (20%) adults with any other primary source of infection.

### Phenotypic antimicrobial susceptibility profile

The phenotypic antimicrobial susceptibility test (AST) profile for the 322 *E. coli* isolates, measured by disc diffusion, is shown in Figure 2. There was a high prevalence of resistance to the β-lactam + β-lactamase inhibitor combination amoxicillin + clavulanic acid (co-amoxiclav), with 191 (59%) isolates being resistant. Resistance to piperacillin + tazobactam, another β-lactam + β-lactamase inhibitor combination, was lower (34 (11%), 21 (6.5%) and 267 (83%) isolates were resistant, intermediate and susceptible, respectively). An ESBL and/or AmpC phenotype was detected in 52 (16%) and 13 (4%) isolates, respectively (with a single isolate generating both phenotypes); no carbapenem resistant organisms were identified. Resistance to aminoglycosides was low: 27 (8.4%), 3 (<1%) and 292 (91%) isolates were resistant, intermediate and susceptible to gentamicin, respectively.

**Figure 2.**
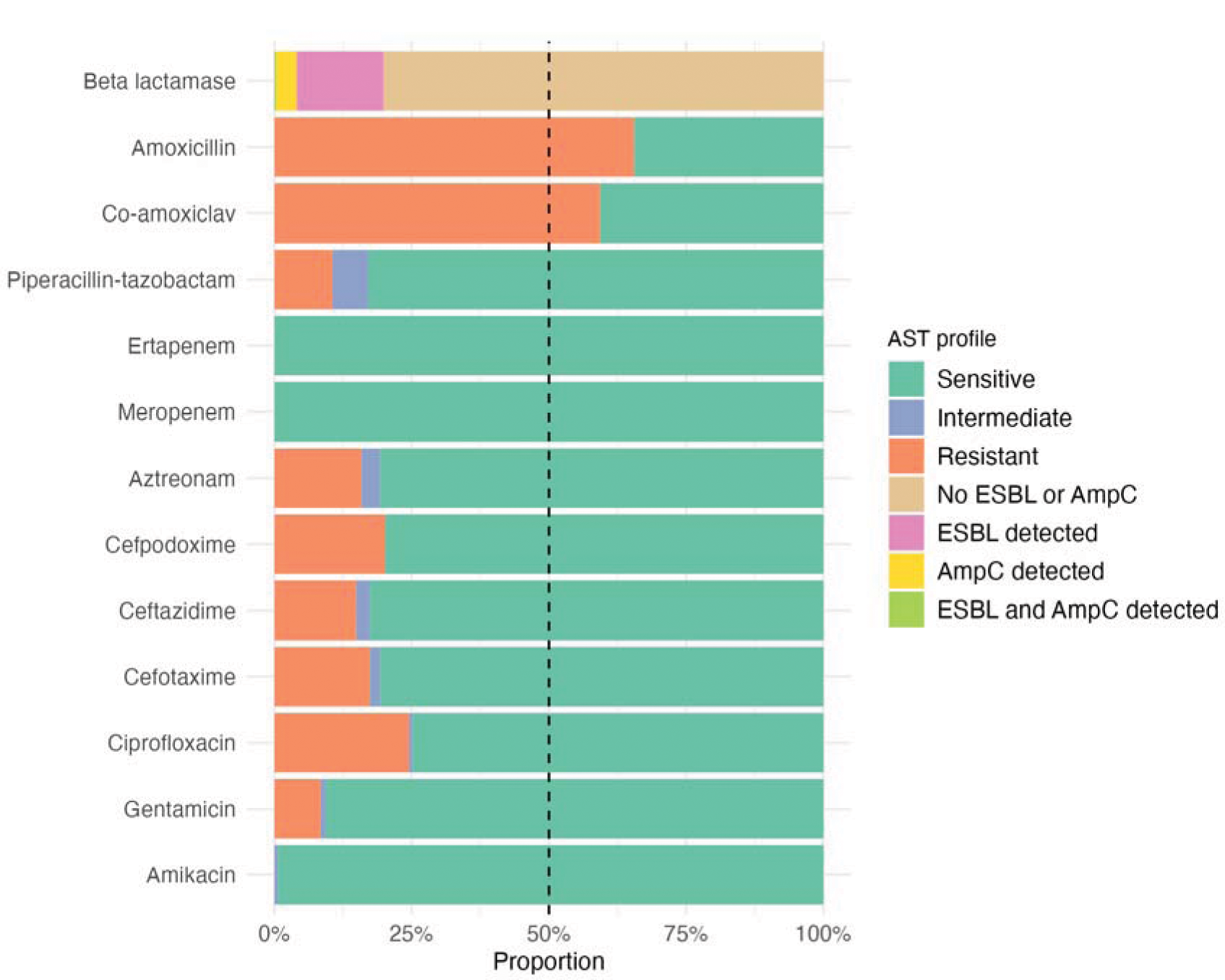
Phenotypic antimicrobial susceptibility profile for all isolates. Antimicrobial susceptibility was determined by disc diffusion with break points interpreted using BSAC zone-size cut-offs. Isolates that were resistant to cefpodoxime were selected for further testing to determine whether an ESBL or AmpC phenotype was present.

Gentamicin is currently the recommended first-line treatment for complicated UTI in CUH; nearly all isolates were susceptible to amikacin (2 (<1%) and 320 (99%) were intermediate and susceptible, respectively).

### Genomic characterisation

The phylogeny of the 322 *E. coli* isolates, generated by the core genome sequences, is shown in Figure 3. In total, 75 sequence types were identified; however, the majority were identified at low frequency, with 42 being represented only by a single sample (Supplementary Table 3, Supplementary Figure 7). Just four STs accounted for over half (54%) of all cases: ST131 (65, 20%), ST73 (51, 16%), ST69 (33, 10%), and ST95 (28, 8.7%). The core genome sequences generated a well-defined population structure, which correlated precisely with assigned ST (i.e., genetic diversity was lower within sequence types than between them) (Supplementary Figures 8-9).

**Figure 3.**
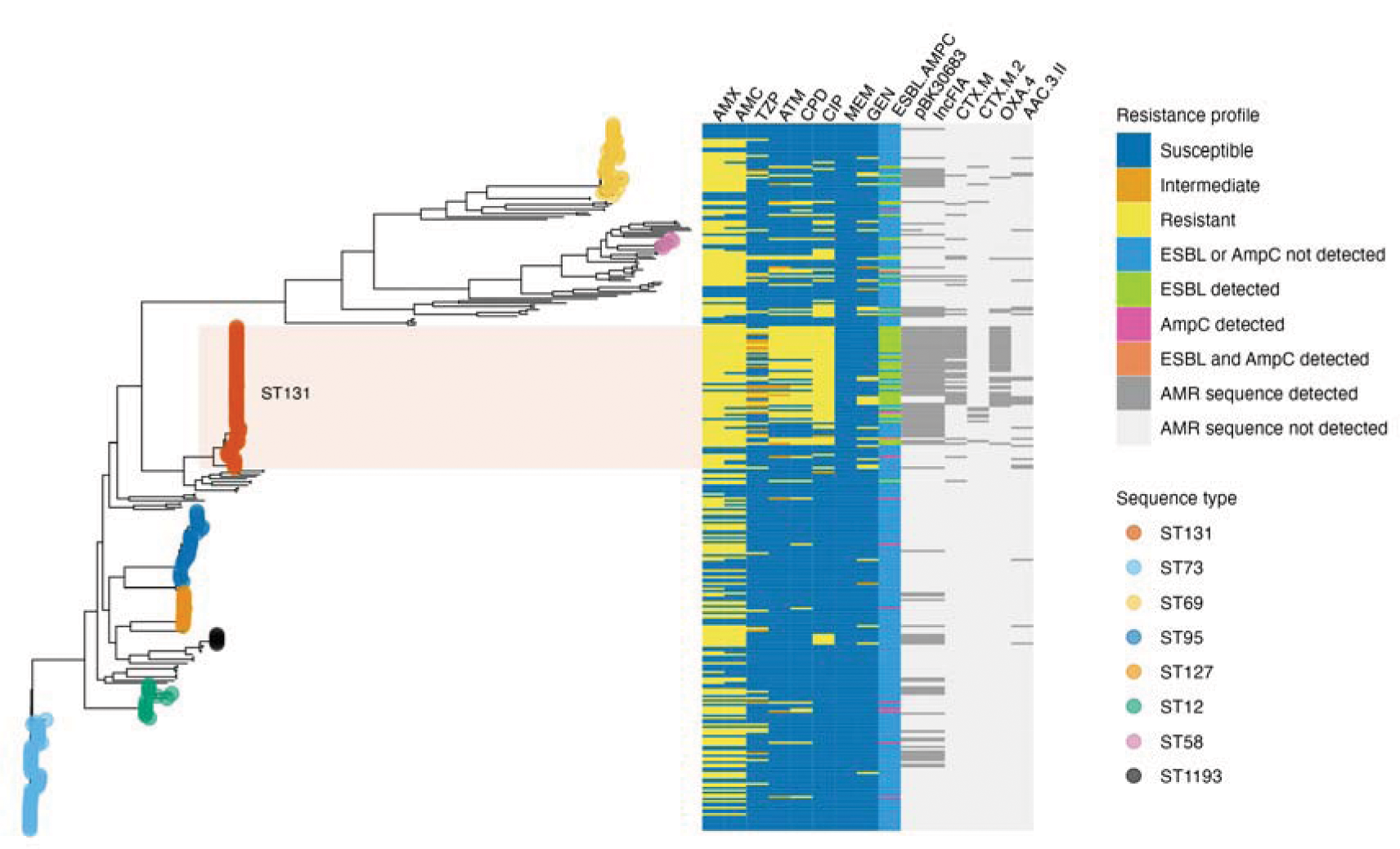
Core genome phylogeny of the *E. coli* study population. Branch tips are coloured by sequence type, showing the 8 sequence types with at least 5 samples. Coloured bars depict phenotypic antimicrobial susceptibility profile (left) and the presence or absence of selected AMR-associated genes and plasmids (right). Sequence type 131 (ST131) is highlighted, which was the most frequent sequence type identified (65/322 (20%) isolates) and associated with high rates of AMR (Figure 4). AMX = Amoxicillin, AMC = Co-amoxiclav, TZP = Tazocin, ATM = Aztreonam, CPD = Cefpodoxime, CIP = Ciprofloxacin, MEM = Meropenem, GEN = Gentamicin, ESBL = extended spectrum β-lactamase, AMPC = AmpC β-lactamase.

ST131 was the most abundant ST and was associated with phenotypic resistance to multiple antimicrobials (Figure 4). Of the ST131 organisms, 37/65 (57%) had an ESBL detected, compared with 15/257 (5.8%) for non-ST131 samples; 51/65 (78%) were ciprofloxacin resistant, compared with 28/257 (11%) for non-ST131 isolates. The unadjusted odds ratio (OR) for ST131 being classified as an ESBL producer and ciprofloxacin resistant was 21 (95% confidence interval, CI, 9.9 - 47, *p*<10, Fisher’s exact test) and 29 (95% CI 14 - 65, *p*<10, Fisher’s exact test), respectively. ST131 isolates were also more likely to possess AMR resistance-associated genes and plasmids, such as CTX-M and OXA-4, and the plasmid structures pBK30683 and IncFIA, than non-ST131 isolates (Figure 3, Supplementary Figure 10).

**Figure 4.**
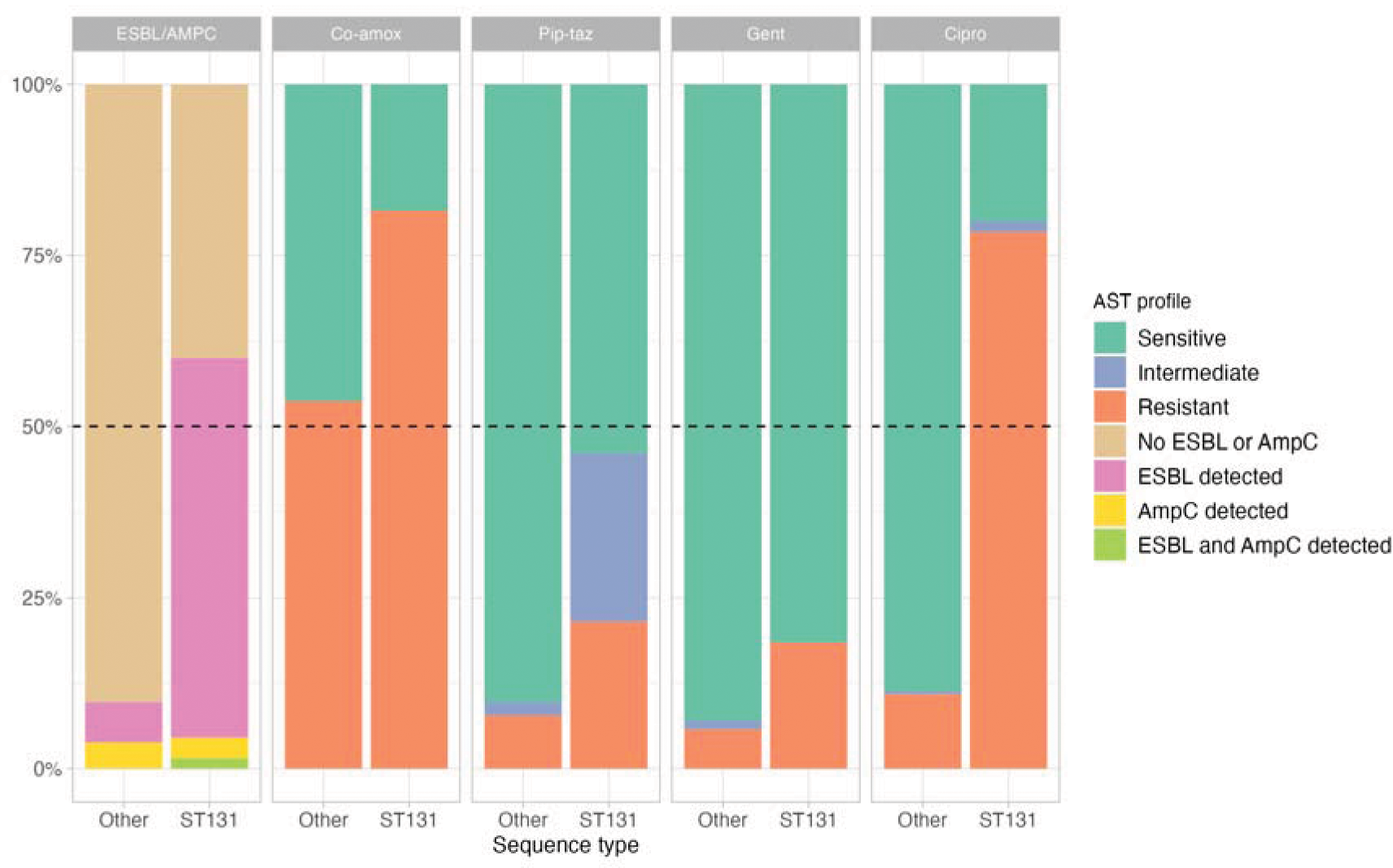
Phenotypic antimicrobial susceptibility profile for selected antimicrobials for ST131 vs non-ST131 isolates. Antimicrobial susceptibility was determined by disc diffusion using BSAC cut-offs for interpretation, and ESBL and AmpC status was determined for cefpodoxime resistant isolates. Isolates are divided into ST131 (n=65) and non-ST131 (n=257). Results for ESBL or AmpC status (“ESBL/AMPC”), amoxicillin-clavulanic acid (“Co-amox”), piperacillin-tazobactam (“Pip-taz”), gentamicin (“Gent”) and ciprofloxacin (“Cipro”) are shown.

### The clinical and epidemiological characteristics of ST131

There was no association between isolates belonging to ST131 and the likelihood for organisms to be healthcare-associated or the source of bacteraemia (Figure 5, Supplementary Figure 11). This observation implies that ST131 can cause invasive disease in both community and healthcare settings. Indeed, all the major STs had a comparable profile of bacteraemia source (most commonly UTI) and the proportion that were healthcare-associated. One exception was ST1193, of which none were associated with UTI; however, this group only contained five isolates.

**Figure 5.**
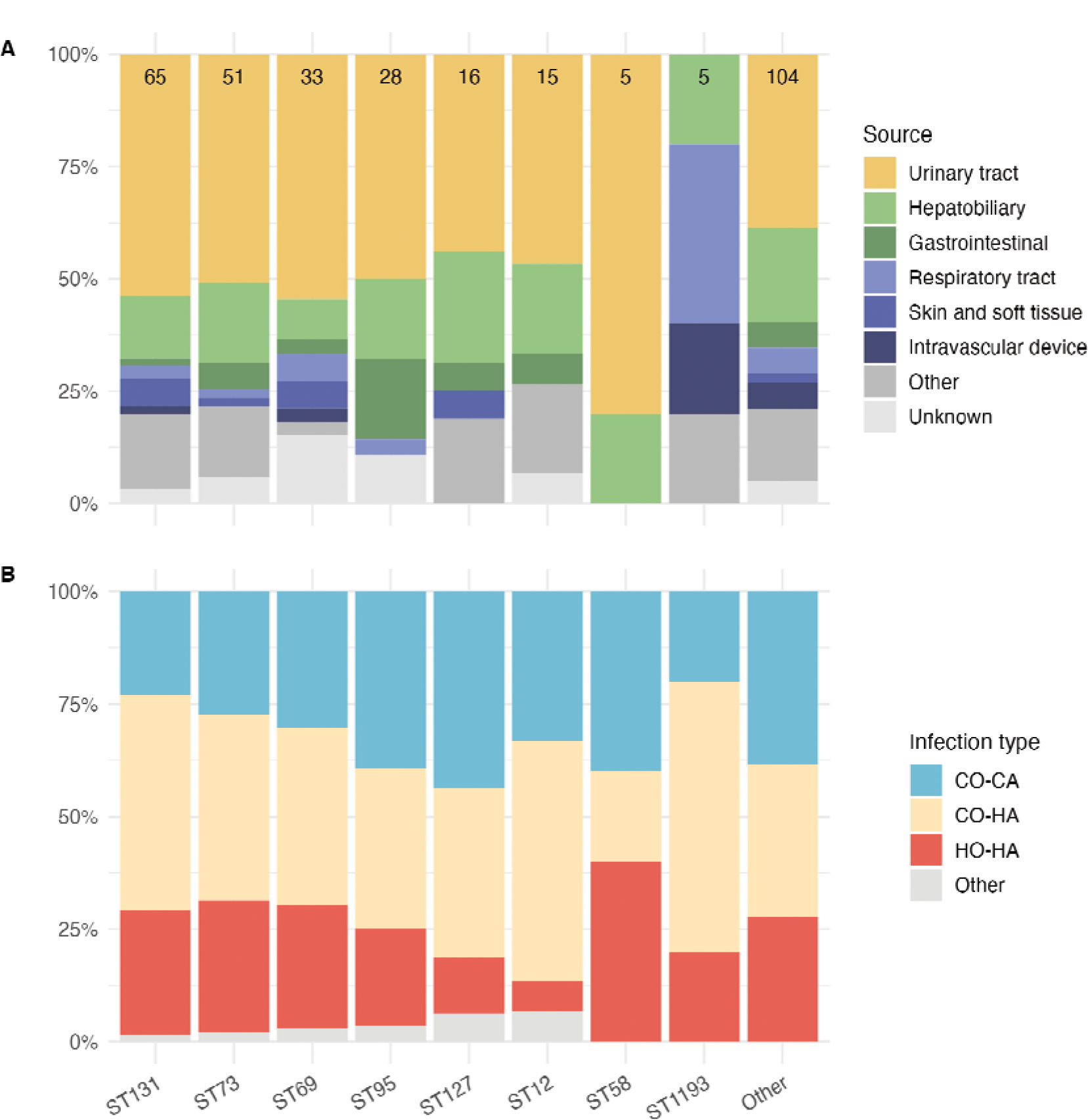
Bacteraemia source and healthcare association class for major sequence types. Showing all sequence types with ≥5 samples, in size order from highest to lowest left to right, followed by all other isolates. Number of isolates in each sequence type displayed in the top row of bars. CO-CA = Community-onset, Community Associated; CO-HA = Community-onset, Healthcare-associated; HO-HA = Hospital-onset, Healthcare-associated.

Disrupting the transmission of AMR within clinical settings is a key goal of infection prevention and control (IPC). We investigated potential epidemiological links between genetically similar ST131 isolates to better understand possible transmission mechanisms. Clusters of highly-similar ST131 isolates were defined with a maximum permitted pairwise SNP difference from the core genome alignment of 17 SNPs within clusters (previously suggested as an epidemiologically relevant cut-off for transmission investigations [41]). We identified two large clusters, referred to as Cluster 1A (11 isolates, with maximum pairwise difference of 12 SNPs) and Cluster 1B (eight isolates, with maximum pairwise difference of 14 SNPs). Both these clusters were entirely comprised of ciprofloxacin-resistant isolates and 17/19 had an ESBL phenotype (Figure 6). We additionally noted a smaller group of MDR organisms, ST1193 (five samples in total), in which 3 samples formed a cluster (with maximum pairwise difference of four SNPs).

**Figure 6.**
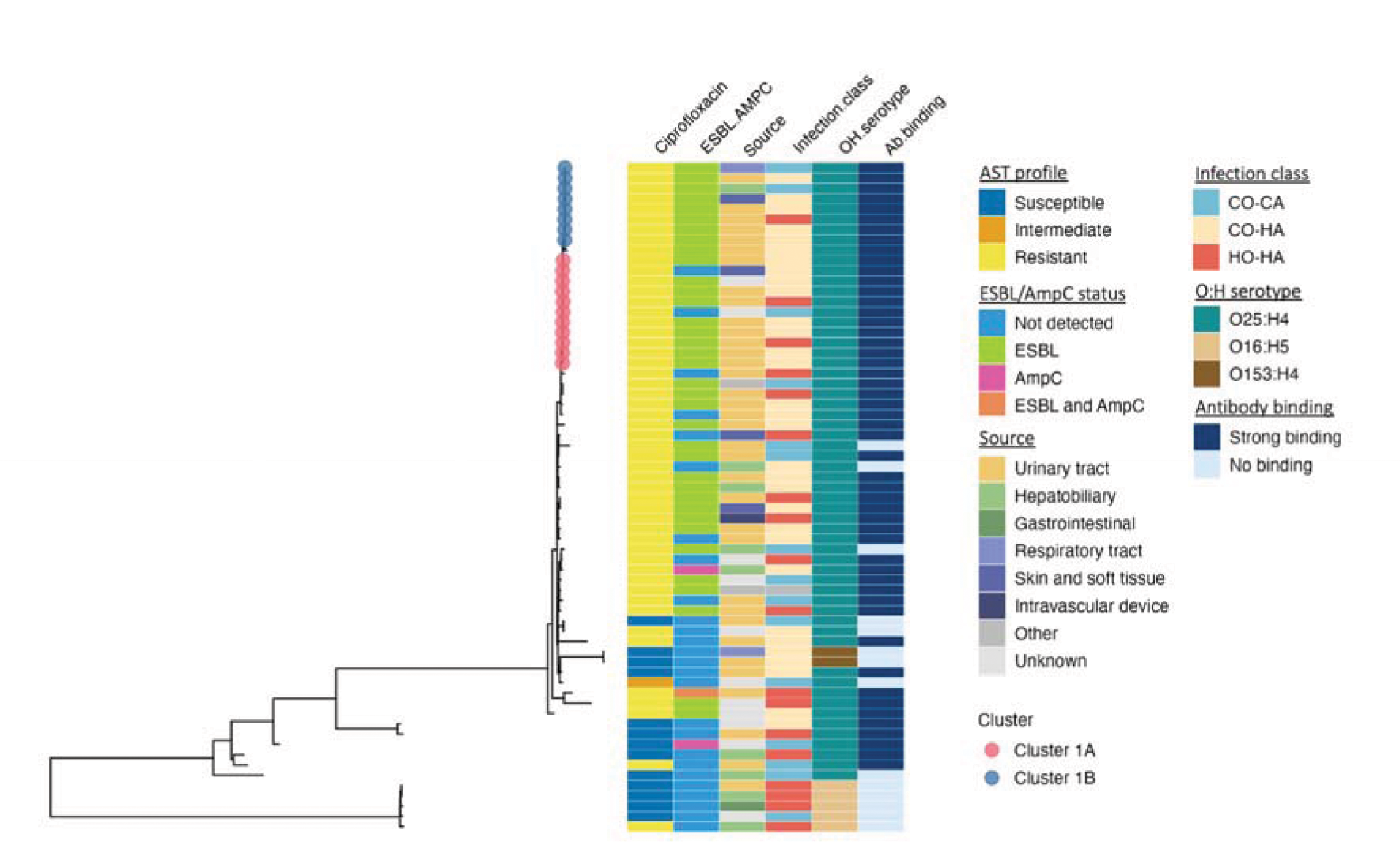
ST131 core genome phylogeny. Sub-tree of the same phylogeny displayed in Figure 3 showing the 65 ST131 isolates. Branch tips depict the two largest clusters of highly similar ST131 isolates. Coloured bars show, from left to right, ciprofloxacin susceptibility (determined by disc diffusion and BSAC interpretations), ESBL or AmpC status (assayed in cefpodoxime resistant isolates), infection source and classification, OH serotype status, and the strength of anti-O25b antibody binding. A large monophyletic group of isolates were nearly all ciprofloxacin resistant, mostly possessed an ESBL, and were nearly all serotype O25:H4 with strong binding to the anti-O25b antibody. As with most *E. coli* isolates, the majority of infections were from a urinary source, and a mixture of community- and healthcare-associated cases were represented. CO-CA = Community-onset, community associated; CO-HA = Community-onset, healthcare-associated; HO-HA = Hospital-onset, healthcare-associated.

Both ST131 clusters included both community- and healthcare-associated cases. On review of patient electronic medical records, no epidemiological links were identified between cases in Cluster 1B or the ST1193 cluster. In Cluster 1A, only weak epidemiological links were identified between two sets of patients. One patient pair overlapped the same ward for a single day but were not placed in the same patient bay. Another pair were placed in the same patient bay but did not temporally overlap. None of the hospital acquired cases showed clinical or epidemiological links to other cases within each cluster. In summary, we did not identify any clear signals of direct patient-to-patient transmission of ST131, even for highly similar isolates.

### O:H serotyping *in silico* and *in vitro*

We evaluated the O:H serotype profile of the isolates *in silico*. 93 O and H serotype combinations were identified (Supplementary Table 4). As with the STs, the distribution was highly skewed; 58 O:H serotypes were represented by only a single sample, while the top five O:H serotypes included 136/322 (42.2%) of all samples. The most common O:H serotype was O25:H4 (60 isolates, 18.6%); which has been associated with ESBL-producing ST131 isolates in several countries. 58/65 (89.2%) ST131 were O25:H4 in this collection, of which 39/58 (67%) had an ESBL and/or AmpC phenotype. The remaining ST131 isolates were serotype O16:H5 (n=5) and O153:H3 (n=2), none of which were ESBL or AmpC producers. The two non-ST131 O25:H4 isolates were both ST95 (neither were ESBL or AmpC producers). The 65 ST131 genomes were aligned against an ST131-specific reference genome and a 347bp fragment of the *pabB* gene was extracted from the alignment (using primers from [42, 43]). This analysis indicated that all O25 isolates from the collection specifically belonged to O25b.

The sensitivity and specificity of an organism being ST131 as a marker of ESBL production in this *E. coli* cohort was 71% and 90%, respectively; and the sensitivity and specificity of ST131 as a marker of ciprofloxacin resistance was 65% and 94%, respectively. The Positive Predictive Value (PPV) and Negative Predictive Value (NPV) an organism being ST131 as a marker of ESBL production was 57% and 94%, respectively, and for ciprofloxacin resistance, of 79% and 89%, respectively. A negative rapid serology test for ST131 in *E. coli* BSI could therefore be an early indicator that an isolate is unlikely to be an ESBL producer; a positive result would suggest the isolate was more likely to be an ESBL producer.

To further explore the potential for an antibody-based rapid diagnostic test for ST131, as a potential marker of ESBL in *E. coli*, we evaluated the binding strength of an anti-O25b antibody against all ST131 isolates. The majority of isolates had strong (49/65, 75%) or strong agglutinating (2/65, 3%) antibody binding (grouped as ‘strong binding’); a minority had no (14/65, 22%) antibody binding (Figure 7). All of the strong binding samples were predicted to be serotype O25b:H4 *in silico*. Of the 14 samples with no binding, 7 were O25b:H4, 5 were O16:H5, and 2 were O153:H4. The clusters of genetically similar, low-diversity *E. coli* ST131 were all O25b:H4 and all exhibited strong antibody binding (Figure 6).

**Figure 7.**
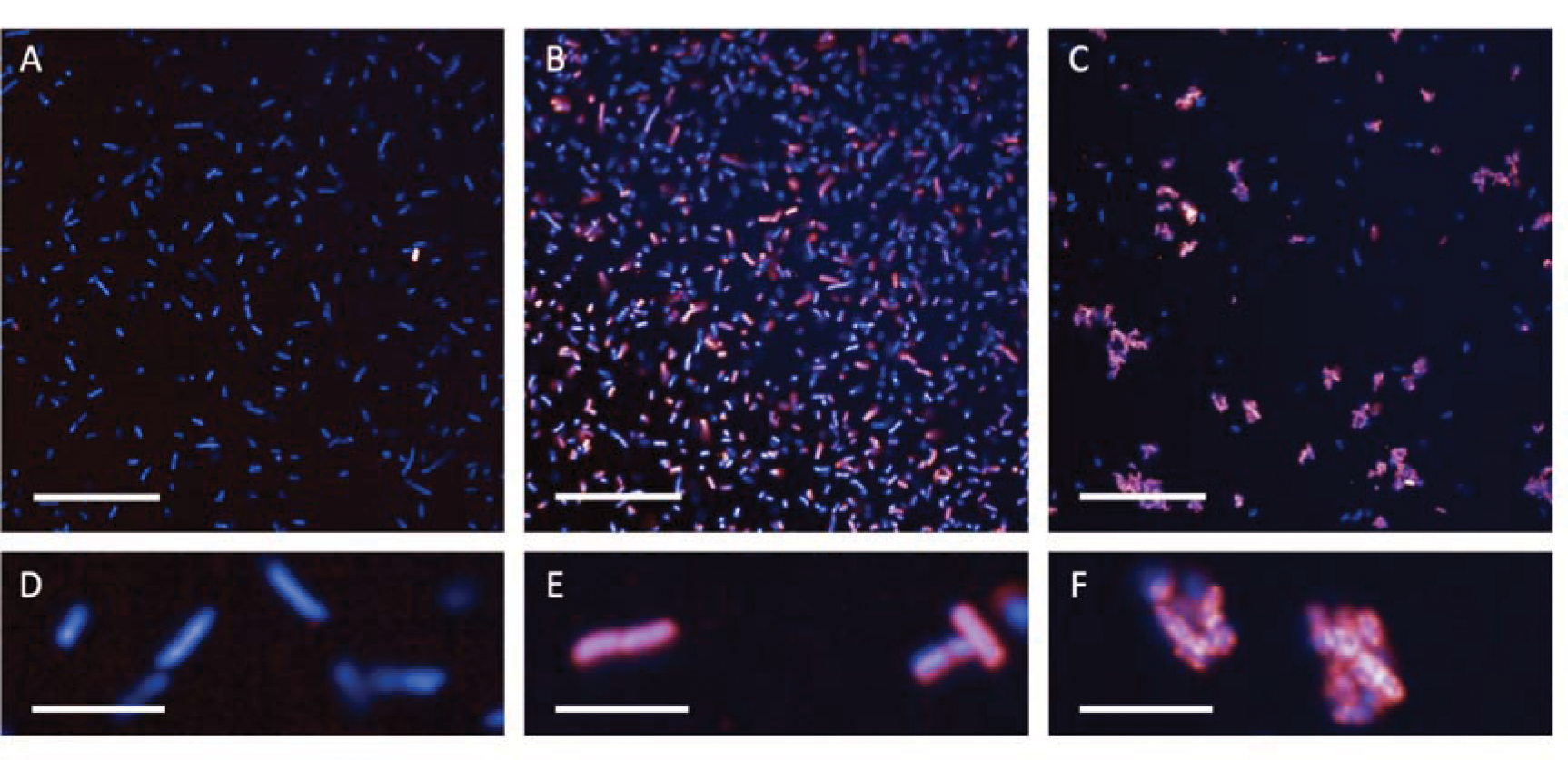
Immunofluorescence of anti-ST131-O25b antibody binding. Shown in blue (DAPI) nucleoid of *E. coli* ST131; shown in red (Alexa-647) KM-467 anti-ST131-O25b antibody. The main phenotypes identified were ‘no binding’ (A, magnified in D), ‘strong binding’ (B, magnified in E), and ‘strong binding with agglutination’ (C, magnified in F). Scale bar: A-C = 50 microns, D-F = 10 microns.

## Discussion

In this study, we combined multiple data sources including clinical, epidemiological, genomic and antibody binding data to characterise *E. coli* BSI in a large UK hospital. Of the 322 isolates included in the study, 12% patients died within 30 days of their bacteraemia, with increasing age and non-urinary source associated with higher odds of death. By disc diffusion testing, 59% isolates were resistant to co-amoxiclav, 16% had an ESBL, and 4% had an AmpC. At the time the samples were collected, co-amoxiclav was the first-line recommended treatment for complex UTI in the hospital; this has since been switched to gentamicin, for which resistance was low (around 90% isolates were susceptible to gentamicin). There were no carbapenem resistant organisms identified.

Consistent with previous work [13], ST131 was the most frequent sequence type, accounting for around 20% of all isolates. The majority of ST131 (around 90%) were serotype O25b:H4, which was associated with ciprofloxacin resistance, ESBL production, and low-diversity clusters. However, these clusters, despite being highly similar across the core genome, generally did not have discernible epidemiological links between them. This raises the question of how genetically similar *E. coli* isolates, including MDR organisms such as clones of ST131-O25b:H4, are spreading between individuals. One possibility is that transmission occurs through contaminated environments, such as fomites. Consistent with this view, ST131 has been found to be prevalent in wastewater samples [44–47]. The organisms may then become incorporated into the human microbiome, periodically causing pathology when such organisms enter a new niche, such as the urinary tract. Another possibility is that transmission occurs in pre-hospital settings such as care homes [12].

ST131, when compared with non-ST131 isolates, was commonly associated with both ciprofloxacin resistance and ESBL production; this observation may have potential for use as an initial rapid screening tool for AMR in *E. coli* BSI in this population. The negative predictive value of ST131 for ciprofloxacin resistance and ESBL illustrates the potential for genomic surveillance combined with antimicrobial susceptibility data to identify markers of clinically salient phenotypes, such as AMR, for the development of rapid diagnostics. Screening tests such as this rely on understanding the relationship between the screening target (e.g. ST131) and the phenotype of interest. For example, an ST131 screening test would lose its high negative predictive value if applied in a population in which ESBLs were common in non-ST131 *E. coli* isolates. Therefore, ongoing surveillance, integrating molecular profiling, clinical epidemiology, and traditional microbiology, is required to ensure that tests and targeted therapies remain reliable, and to identify potential new targets as organisms continually evolve.

We acknowledge several limitations to our study. First, it was a single centre study conducted over one year. Sampling longitudinally over several years would allow investigation of evolutionary trends in *E. coli* populations, for example whether ST131 is increasing in frequency relative to other sequence types over time, and how new lineages emerge and spread. However, our findings are consistent with other studies [13], which also showed a small number of sequence types, including ST131, cause the majority of invasive disease. Second, we did not sample from the community to contextualise the BSI *E. coli* genomic diversity; for example, within the gut microbiome or in uncomplicated lower UTI managed in the community. Wastewater sampling suggests that the genomic profile of invasive *E. coli* is highly skewed compared with the commensal *E. coli* population present in the human gut [47]. Finally, there were limitations on what clinical metadata could be retrospectively collected. For example, we were unable to reliably identify whether UTI were catheter-associated, particularly if there had been catheter manipulation in the community prior to admission to hospital. This was often poorly documented in the electronic medical records, and may represent an opportunity for improved documentation for Gram-negative bacteraemia surveillance going forward.

In summary, we present the clinical, epidemiological, genomic and antibody-binding profile of a cohort of over 300 invasive *E. coli* isolates, identifying clusters of MDR ST131-O25b:H4 organisms that are maintained in both healthcare-associated and community settings. Reducing the spread of these MDR clusters should be a key target for combating AMR. Antigenic correlates of virulence and AMR in a population, such as O25b:H4, could be further investigated for potential vaccines, therapeutics and rapid diagnostic tests.

## Data Availability

Genomic data will be released at time of publication.

## Acknowledgements

We are grateful for the assistance of Josefin Bartholdson Scott and Sally Forrest in isolate processing and DNA extraction for this study. We would also like to thank the Clinical Microbiology and Public Health Laboratory staff for their efforts in processing and storing isolates; the Wellcome Sanger Institute (WSI) sequencing team for *E. coli* whole genome sequencing; and the WSI PAM Informatics team for bioinformatics support.

This research was funded by the National Institute for Health Research Cambridge Biomedical Research Centre at the Cambridge University Hospitals NHS Foundation Trust. The views expressed are those of the authors and not necessarily those of the NHS, the NIHR or the Department of Health and Social Care. This work uses data provided by patients and collected by the NHS as part of their care and support.

