## Supplementary materials for "The clinical, genomic, and microbiological profile of invasive multi-drug resistant Escherichia coli in a major teaching hospital in the United Kingdom"

#### Title

### Supplementary methods

#### Bacterial DNA extraction

All isolates were streaked from frozen stocks onto Iso-Sensitest or Luria Bertani (LB) agar (Oxoid), and incubated overnight at 37°C in a static incubator. Single colonies were picked into a single well of a 96 deep well plate containing Iso-Sensitest broth, before overnight incubation. Bacteria were pelleted and re-suspended in PBS and 20 µL of 100 mg/mL lysozyme solution. DNA extraction was performed using either the MagNAPure small volume nucleic acid extraction kit (Roche) on the FLOW flex platform (Roche), or using the QIAamp 96 DNA QIAcube HT Kit (Qiagen), according to the manufacturer's instructions. Resultant DNA concentration and purity was quantified using a NanoDrop spectrophotometer (Thermo Fisher Scientific) and a Tapestation 4200 using D1000 ScreenTapes (Agilent Technologies).

#### Bioinformatics

Command line used for *roary* core genome alignment:

roary -e -n -cd 99 -p 8 ../*.gff

Command line used for producing phylogenetic tree from the core genome alignment using *iqtree*:

iqtree -s core_genome_alignment.aln -m GTR+G -bb 1000

Command line for producing SNP distance matrix from the *roary* core genome alignment:

snp-dists core_genome_alignment.aln > core_genome_snp_dists.csv -c

Command lines using *ariba* to prepare the CARD database (Version 3.1.1, downloaded from <https://card.mcmaster.ca/>):

ariba getref card out.card

ariba prepareref -f out.card.fa -m out.card.tsv prepareref.out.card

A generic version of the command line used to analyse the samples using the prepared CARD database is shown below, which was looped through all samples:

ariba run prepareref.out.card/ LANEID/LANEID_1.fastq.gz LANEID/LANEID_2.fastq.gz LANEID_out.card

The *ariba* output was summarised as below (generic version of command line):

ariba summary out.card.summary LANEID1_out.card/report.tsv LANEIDn_out.card/report.tsv

*plasmidfinder* was used to identify plasmids. The *plasmidfinder* database was accessed using:

        	ariba getref plasmidfinder out.plasmidfinder

The *plasmdfinder* database was prepared using *ariba*:

ariba prepareref -f out.plasmidfinder.fa -m out.plasmidfinder.tsv prepareref.out.plasmidfinder

*E. coli* MLST alleles were retrieved using:

        	getmlst.py --species 'Escherichia coli#1

Where the *E. coli* MLST fasta file used was Escherichia_coli#1.fasta. MLST typing was performed using *srst2*, example command below (looped through each sample):

srst2 --input_pe strainA_1.fastq.gz strainA_2.fastq.gz --output strainA_test --log --mlst_db Escherichia_coli#1.fasta --mlst_definitions profiles_csv --mlst_delimiter _

And outputs compiled:

srst2 --prev_output *__results.txt --output ecoli_mlst_report

*In silico* OH serotyping was also performed using *srst2*. Example command line used:

srst2 --input_pe LANEID/LANEID_1.fastq.gz LANEID/LANEID_2.fastq.gz --output OH_serotypes --log --gene_db [PATH]/EcOH.fasta

And the data compiled:

srst2 --prev_output *__results.txt --output ecoli_oh_serotype_report

In addition, the 65 genomes identified as ST131 using the *srst2* method were mapped against the ST131 reference genome, Escherichia_coli_UPEC_ST131_v0.2. Primers for the *pabB* gene (TCCAGCAGGTGCTGGATCGT and GCGAAATTTTTCGCCGTACTGT) were used for *in silico* PCR for O antigen typing, taken from [1] and [2]. The 347bp amplicon locus was retrieved from the whole genome alignment (positions 2143355 – 2143701 from the reference genome) and inspected in *AliView*.

#### Categorisation of *E. coli* infection class

Classifications were adapted from mandatory reporting surveillance guidance from UKHSA: <https://assets.publishing.service.gov.uk/government/uploads/system/uploads/attachment_data/file/859759/HCAIDCS_Gram_negative_Submission_Form_2020.pdf>

**Health-care onset** was defined as blood culture positive more than 48 hours after admission. **Health-care associated** was defined as blood culture positive less than 48 hours after admission and one of more of the following criteria: hospital admission in previous 28 days, chemotherapy or neutropenia (less than 0.5) in previous 28 days, surgery in previous 28 days, antibiotic prescription in community in previous 28 days, urinary catheter insertion or manipulation in 28 days before admission. **Community onset** was defined as blood culture positive less than 48 hours after admission and nil of above risk factors identified from review of clinical note and GP entries in month before admission.

#### ST131 antibody binding assay

Bacteria were cultured from storage beads onto LB agar plates. Single colonies were picked and cultured in LB broth overnight at 37⁰C at 200 RPM. Four additional bacterial isolates known from previous work to have different binding patterns were used as controls.  Overnight bacterial cultures were diluted 1:200 in LB broth and 50µl of this bacterial suspension was added per well in CellCarrier-96 Ultra plates (Perkin Elmer) and left in a static incubator for 2 hours at 37⁰C. The supernatant was discarded and any remaining bacteria fixed with 4% paraformaldehyde (PFA) in phosphate buffered saline (PBS) for 15 minutes at room temperature. After washing with PBS, 100µl of the antibody KM467 (diluted to 1µg/ml in PBS + 1% bovine serum albumin) was added for 1 hour at room temperature. After further wash with PBS, 100µl of 2µg/ml Alexa Fluor 647 Goat Anti-Human IgG (ThermoFisher) plus 2µg/ml DAPI (Thermo Fisher) in PBS + 1% bovine serum albumin were added and the plate was incubated for 30 minutes in the dark. After a final wash with PBS, 50µl of PBS was added to each well and the plate was taken for imaging. The antibody binding assay was repeated fivefold.

### Supplementary figures

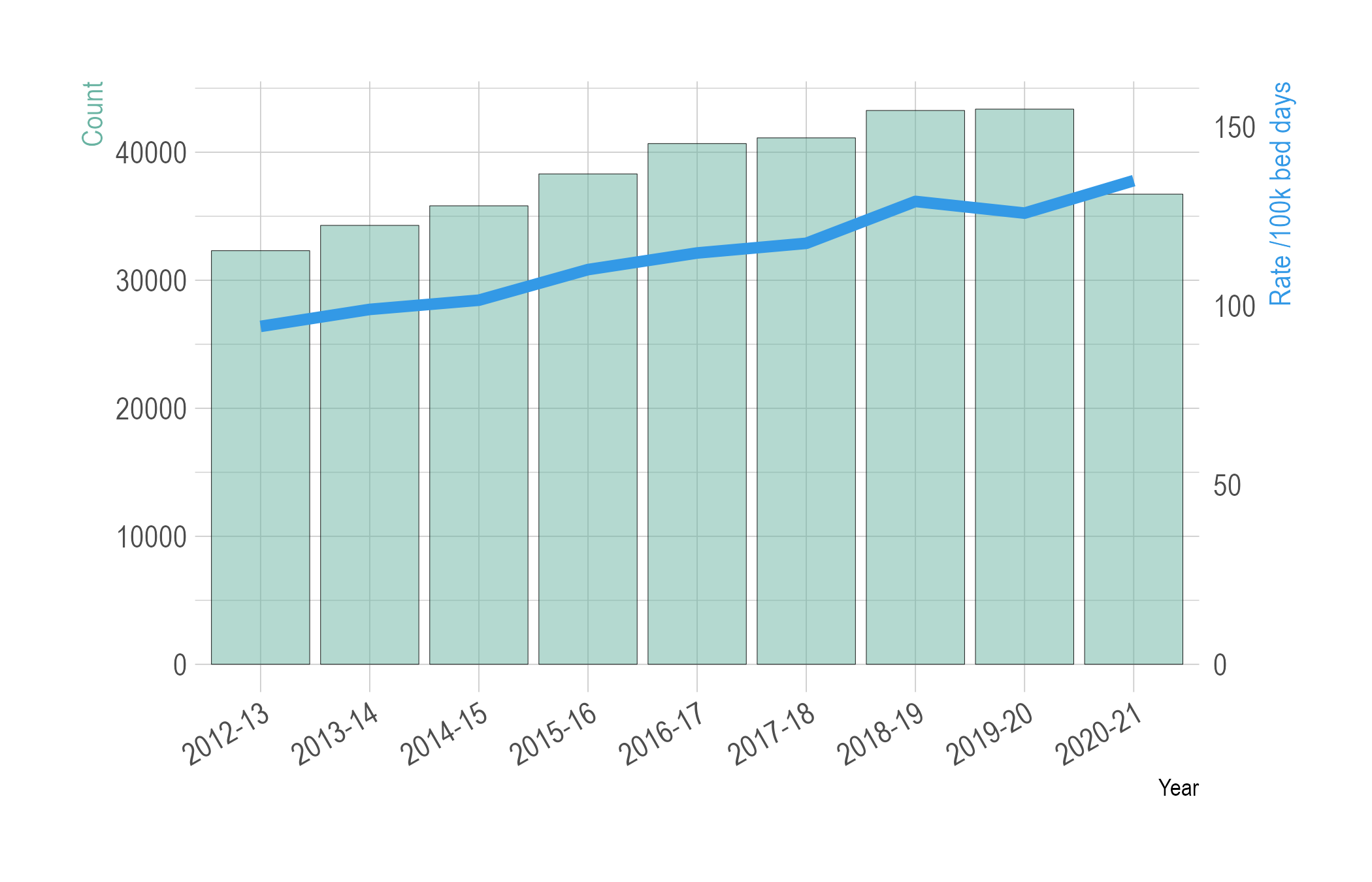

**Supplementary Figure 1.** *E. coli* bacteraemias in NHS acute Trusts 2012-2021. Plotted from data from UK-HSA, ‘Escherichia coli bacteraemia: annual data’, URL: <https://www.gov.uk/government/statistics/escherichia-coli-e-coli-bacteraemia-annual-data>, accessed 2023-02-27.

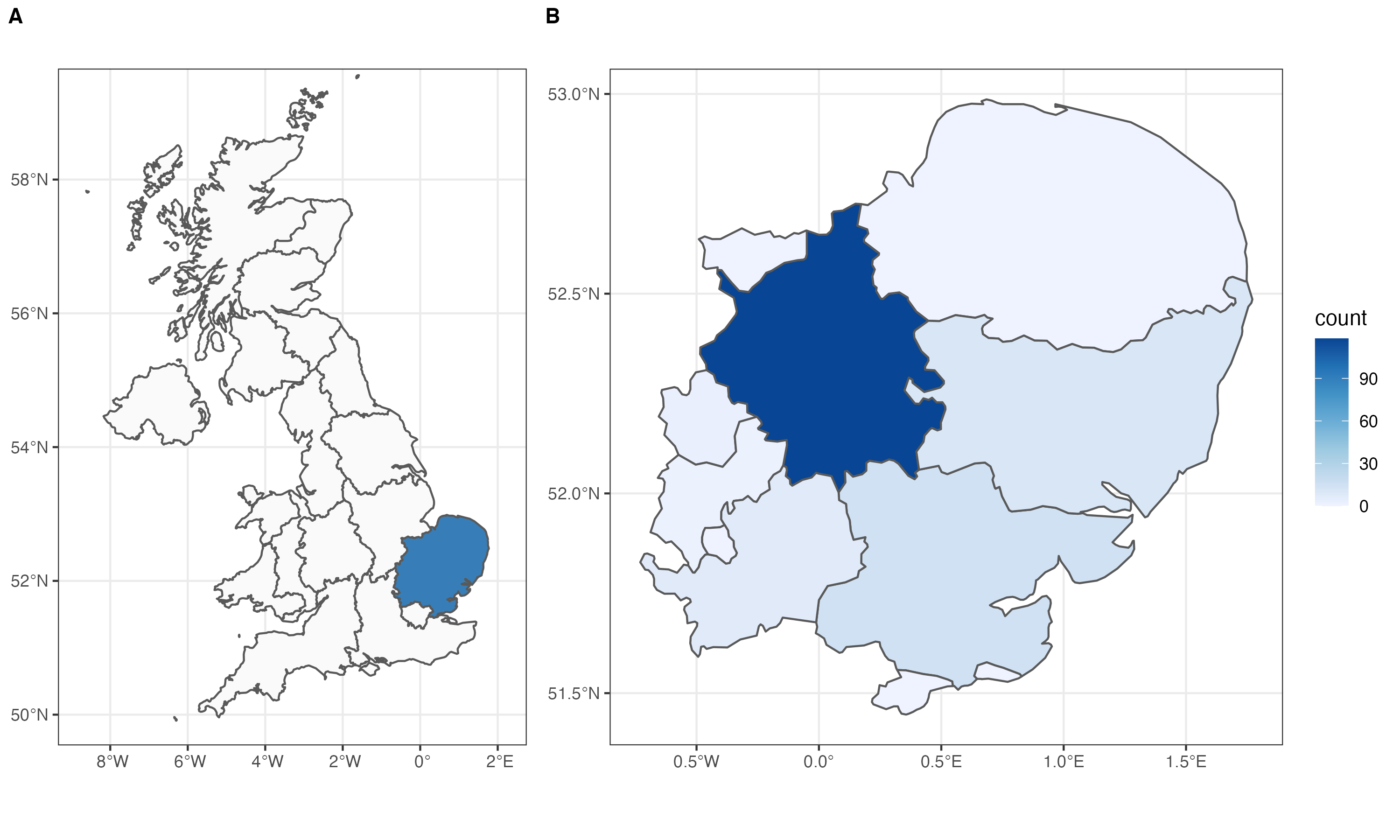

**Supplementary Figure 2.** Study location in the United Kingdom (UK) and geographic distribution of E. coli samples. A) Map of the UK with East of England region highlighted in blue. B) Heat map of East of England coloured by sample count used in this study, for the 161 samples in which patient address information was available. 73% of cases were from Cambridgeshire.

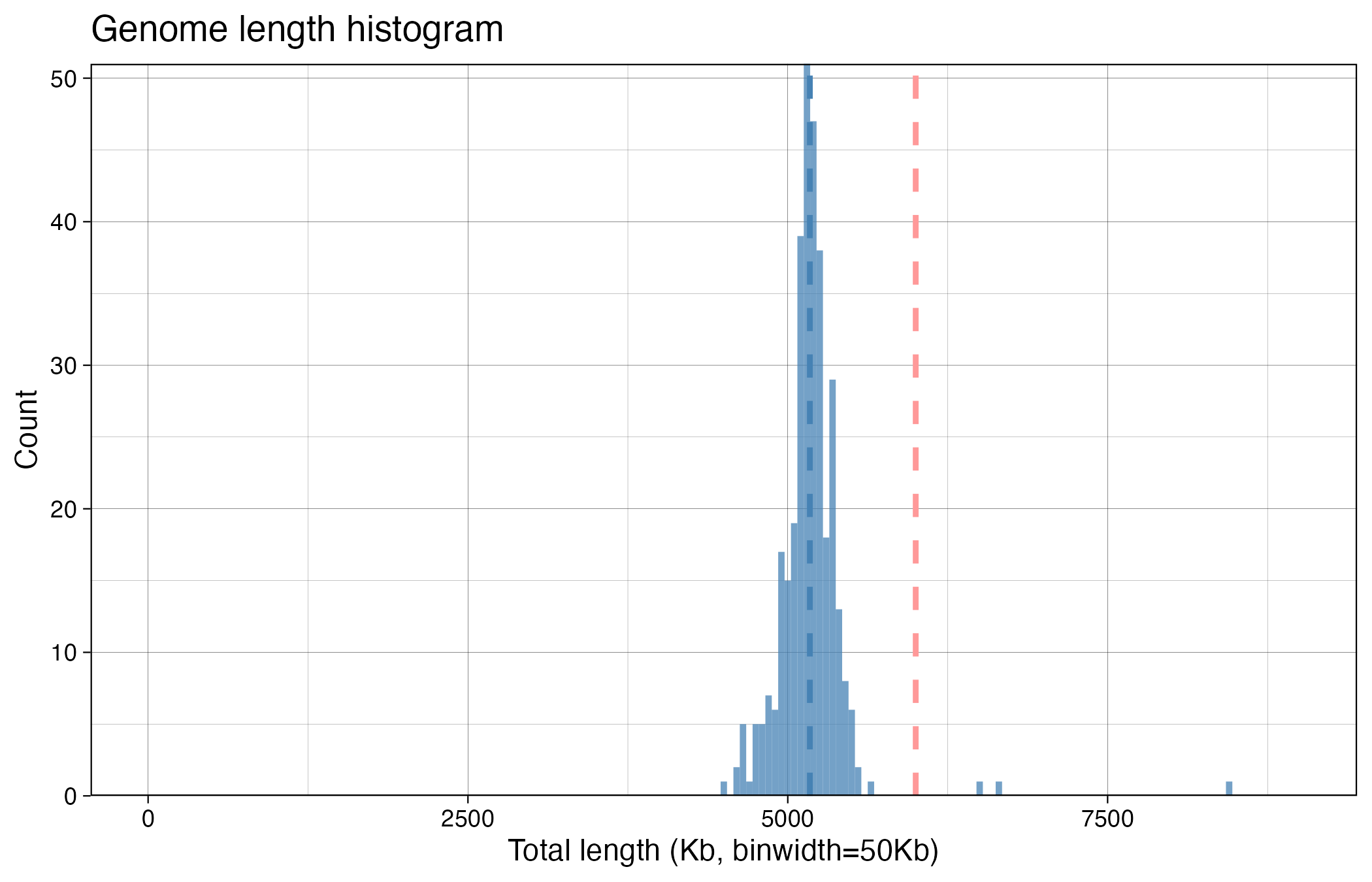

**Supplementary Figure 3.** Histogram of bacterial genome length for the 338 samples pre-QC filtering. Dotted vertical lines show median (blue) and QC cut-off (<6MB, red).

 
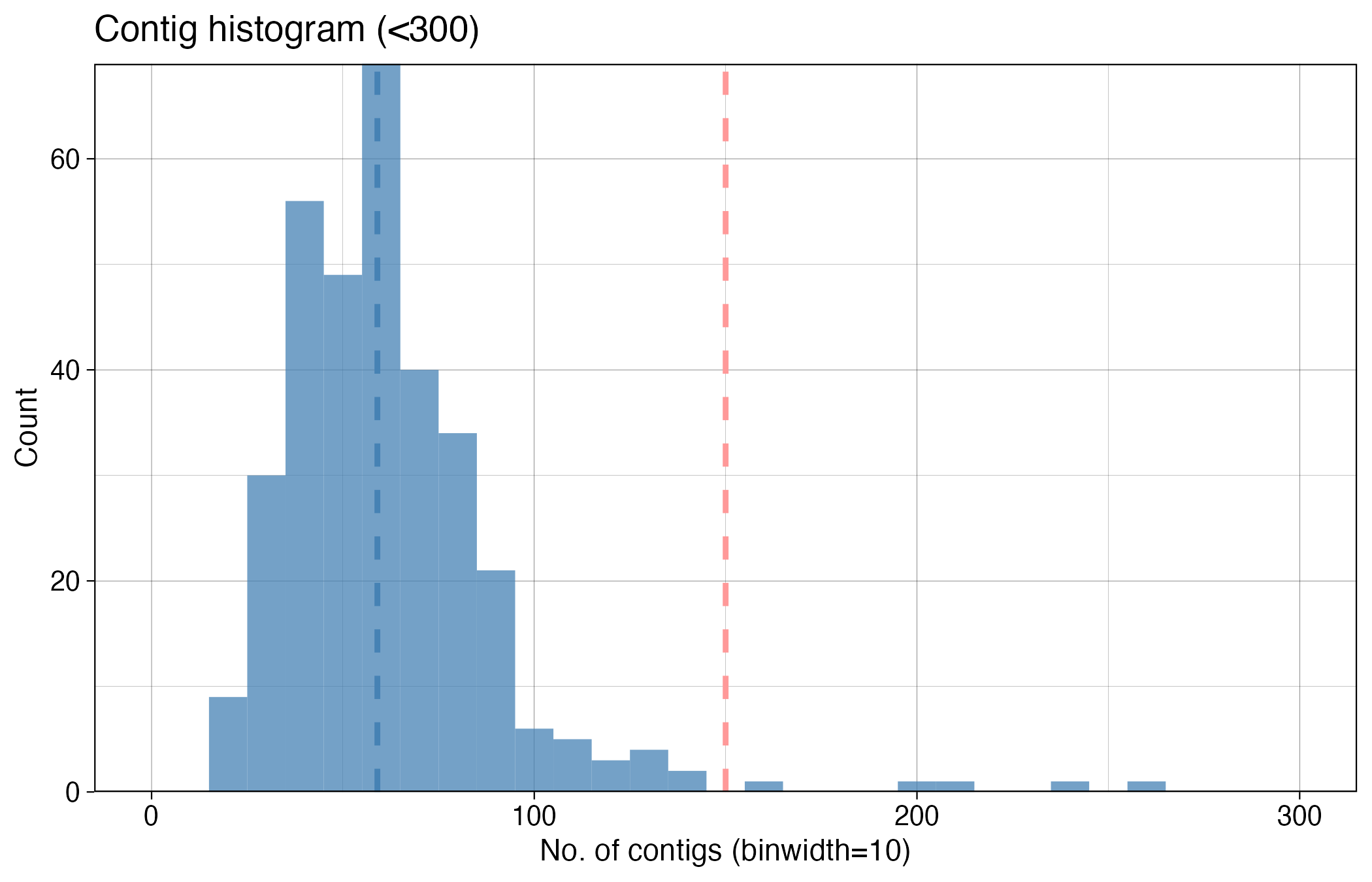

**Supplementary Figure 4.** Distribution of number of contigs per sample for the 338 samples pre-QC filtering. 5 outliers with >300 contigs not shown. Dotted vertical lines show median (blue) and QC cut-off (<150 contigs, red).

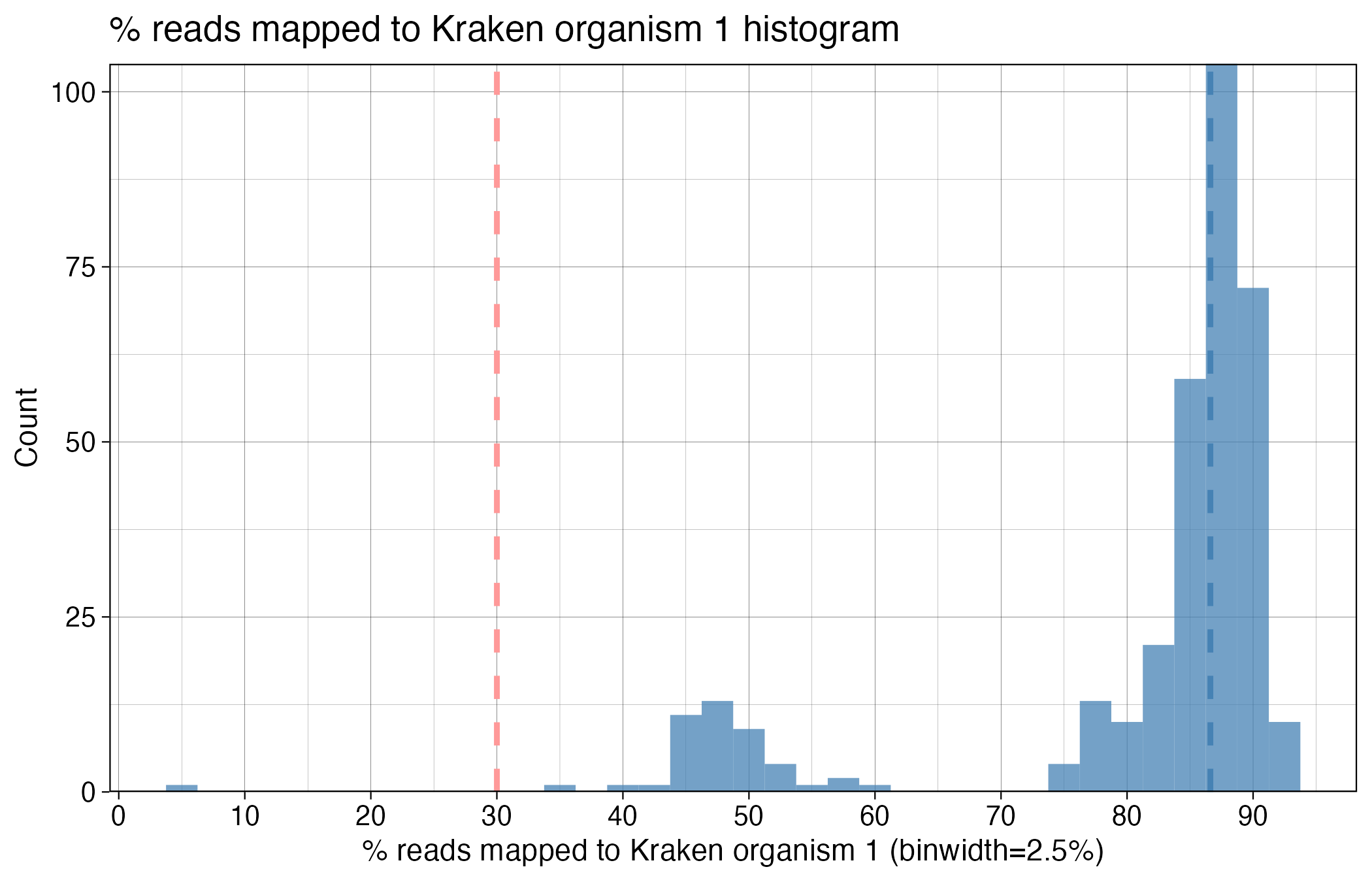

**Supplementary Figure 5.** Histogram of % reads mapping to Kraken organism 1 for the 338 samples pre-QC filtering. Dotted vertical lines show median (blue) and QC cut-off (>30%, red).

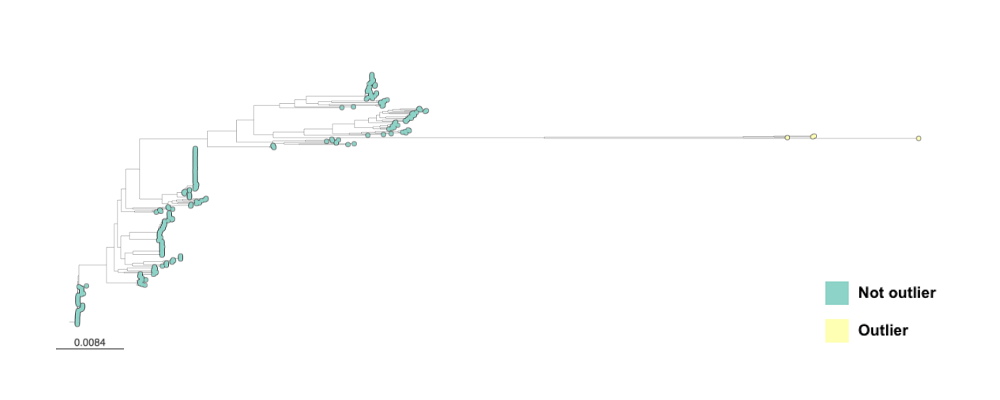

**Supplementary Figure 6.** Phylogenetic tree of 326 samples following initial QC filtering. Core genome alignment performed using *roary* (requiring genes to be present in 99% samples), followed by tree generation from the core alignment file using IQTREE as described in Methods. Four samples were phylogenetic outliers (CUH_ECOL0134, CUH_ECOL0160, CUH_ECOL0400, CUH_ECOL0481 - coloured pale yellow on the tree). The four samples had relatively low % reads mapping to kraken 1 (*E. coli*), ranging from 38% for CUH_ECOL0134 (the lowest of the 326 samples included post-QC) to 47% for CUH_ECOL0400. In all four cases, kraken 2 was *Salmonella enterica*. It is possible these samples are mixed with non-*E. coli* organisms, causing the long branch. The four outliers were removed to yield the final analysis set of 322 samples described in the paper main text. Image downloaded from microreact.

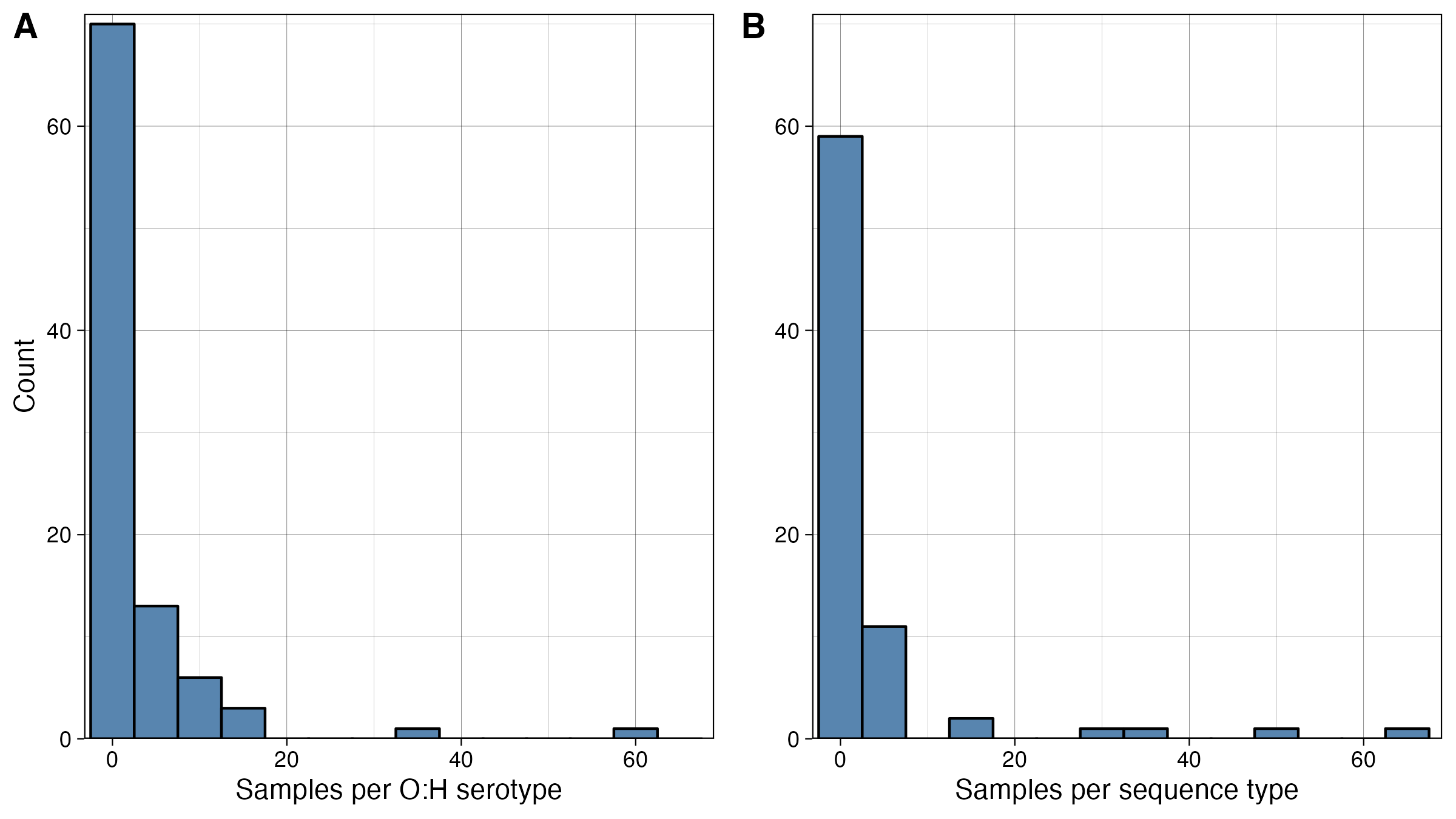

**Supplementary Figure 7.** Histograms of O:H serotype (A) and sequence type (B) frequencies for the 322 *E. coli* genomes in the study. Histogram bin width = 5 for both plots. A small minority of both O:H serotypes and sequence types include many samples – the most frequent O:H serotype, O25:H4, includes 60 (18.6%) samples, while 58 O:H serotypes are only represented by a single sample. Similarly, the most frequent sequence type, ST131, includes 65 (20.2%) samples, while 42 sequence types are only represented by a single sample.

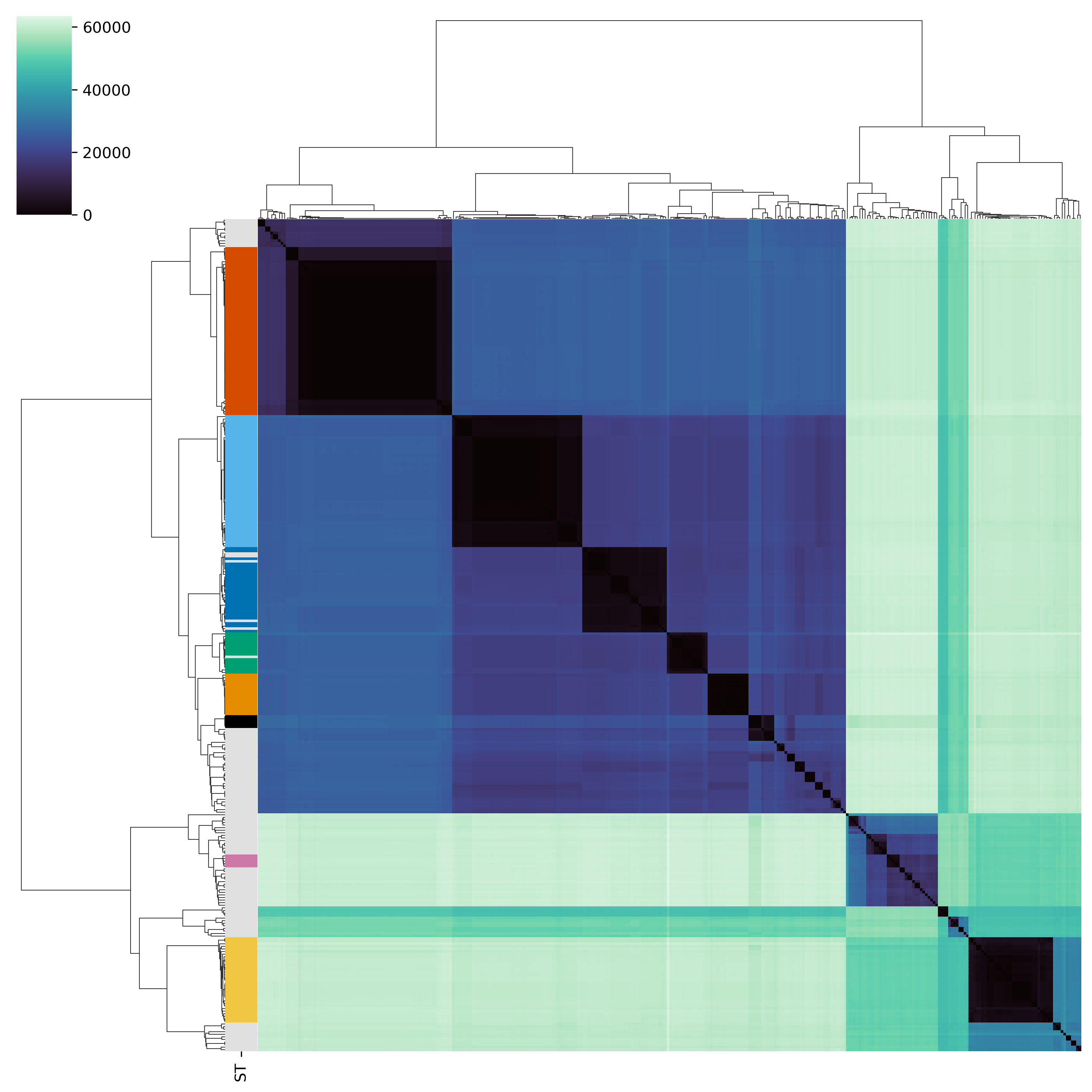

**Supplementary Figure 8.** SNP difference distance matrix highlighting major sequence types (coloured bar, right-hand side). There were low pairwise SNP difference counts between samples within each sequence type. SNP differences were counted from the core genome alignment (produced using *roary*), using *snp-dists*.

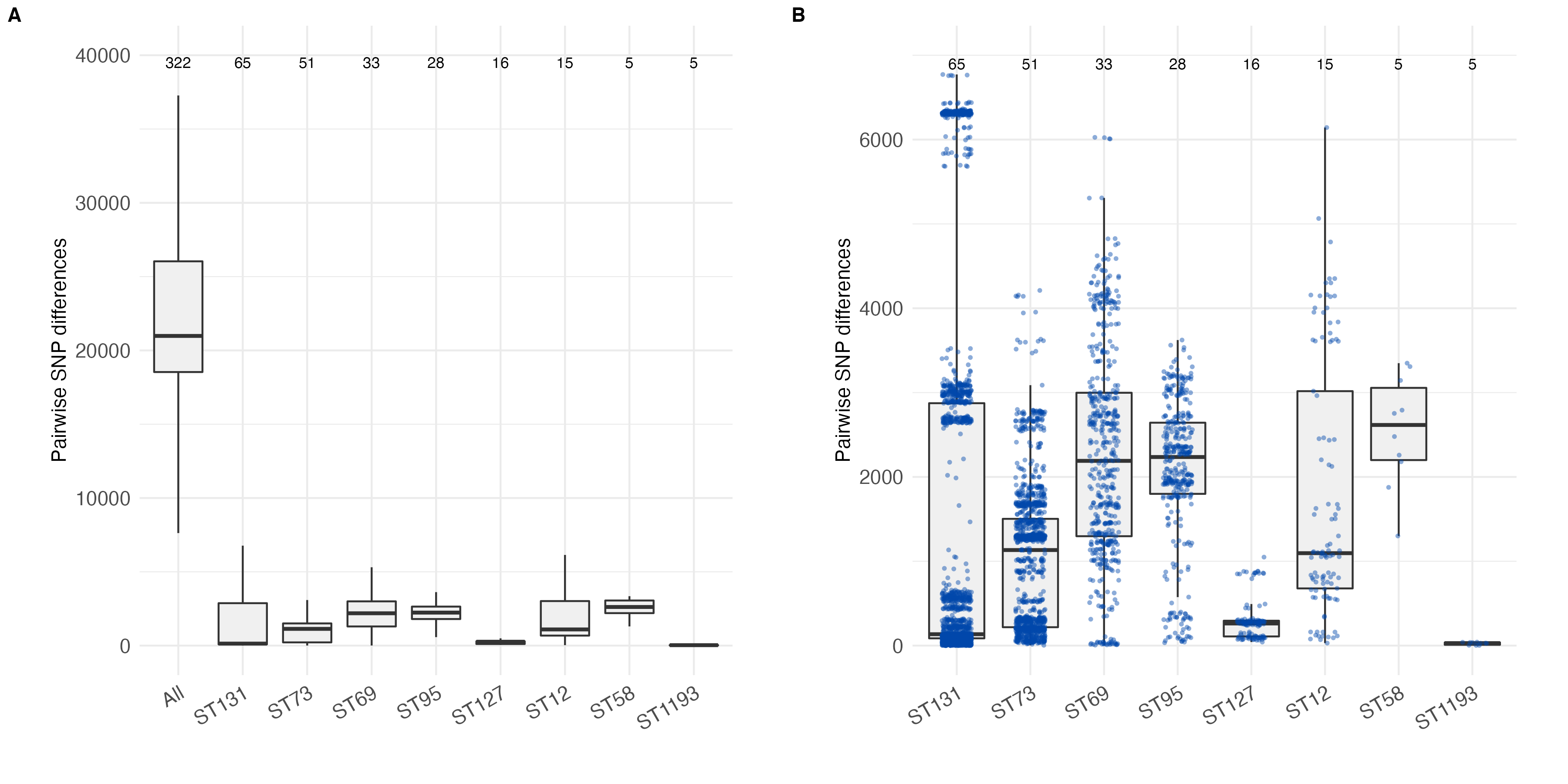

**Supplementary Figure 9.** Pairwise SNP differences between isolates for different samples grouped by Sequence Type (ST).

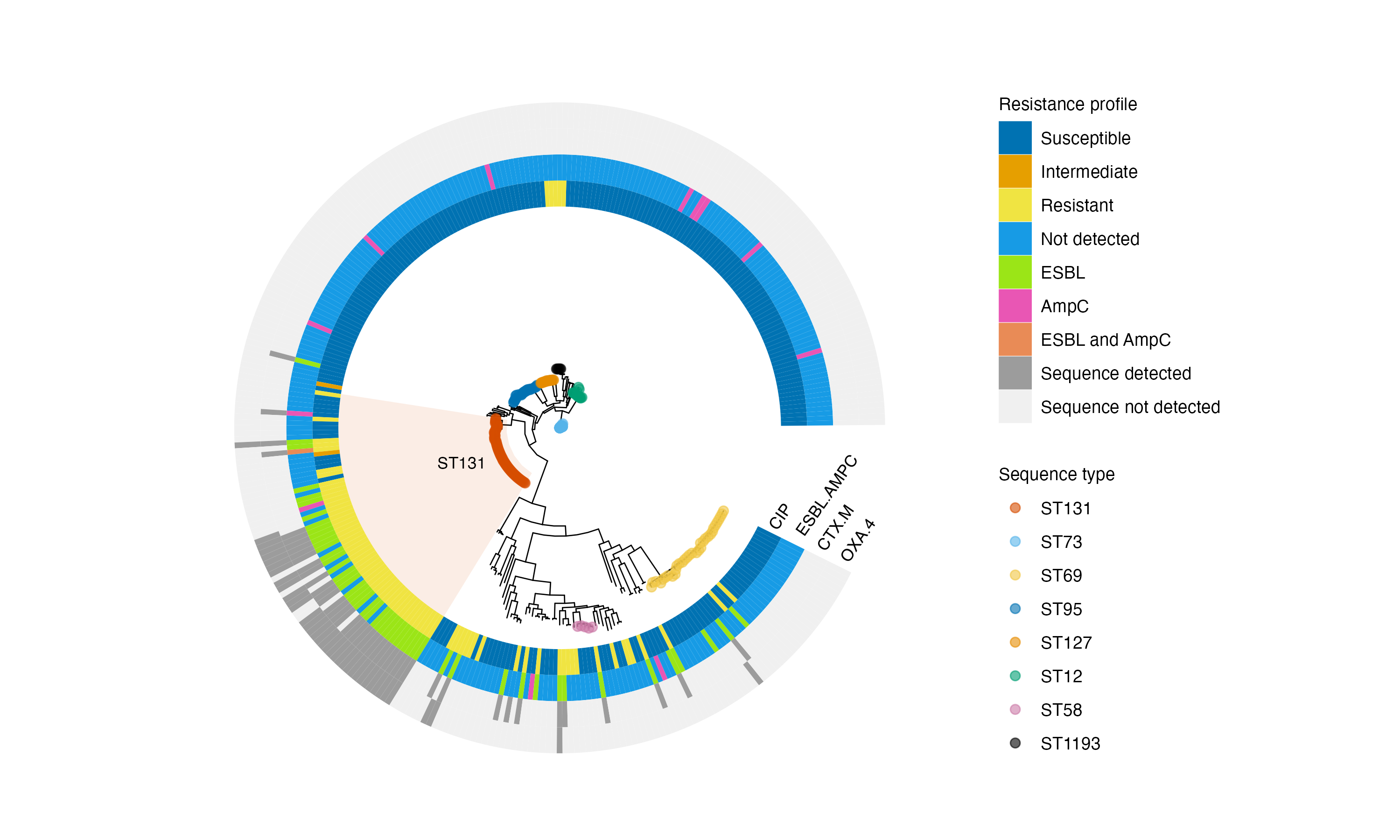

**Supplementary Figure 10.** Core genome phylogeny highlighting sequence type 131, which was associated with ESBL production and ciprofloxacin resistance phenotypically, and high rates of CTX-M and OXA-4 detection genetically. CIP = ciprofloxacin.

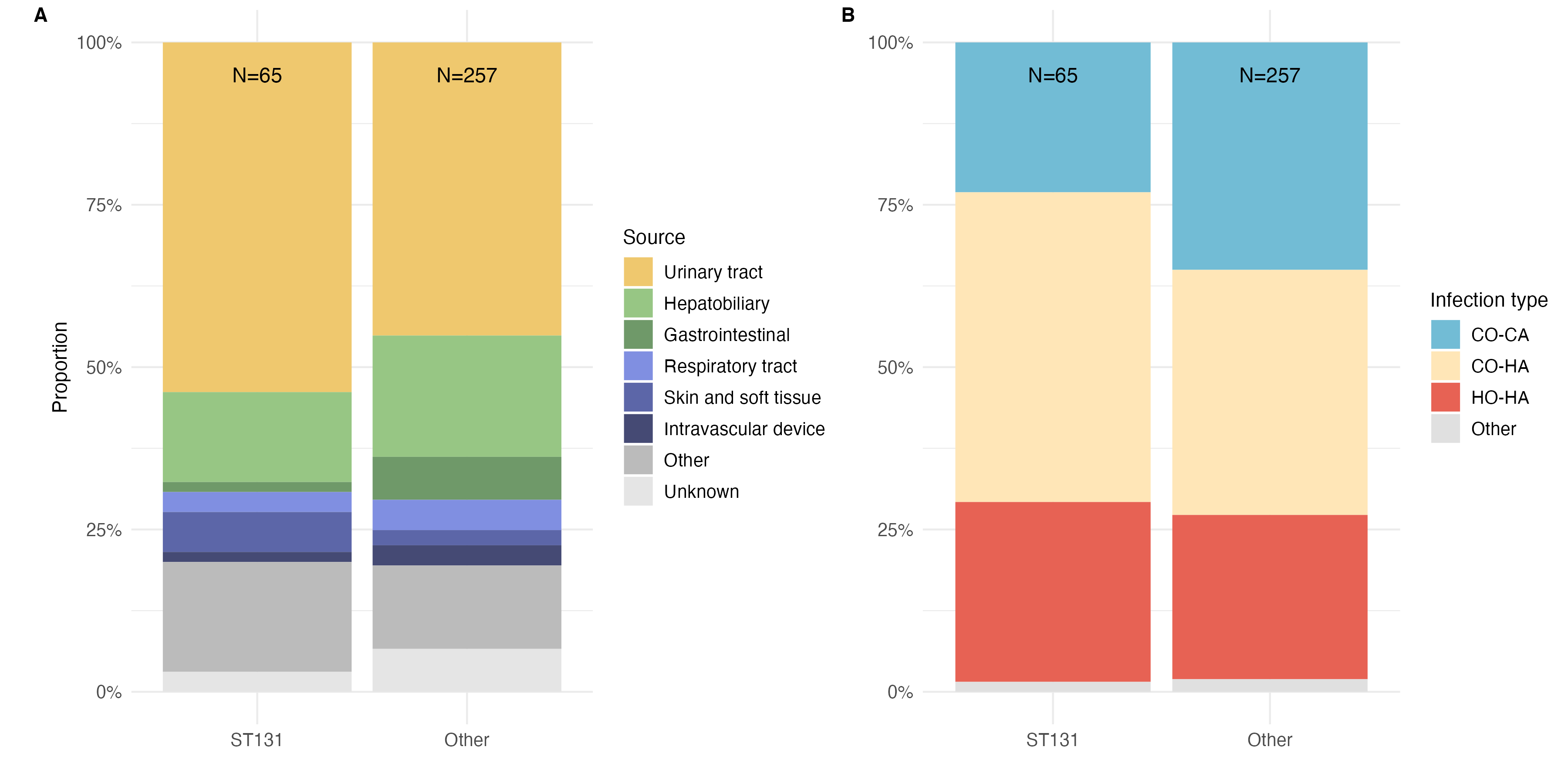

**Supplementary Figure 11.** Bacteraemia source and infection class for ST131 vs non-ST131 isolates.

### Supplementary tables

|  | **Description** | **Counts** |
| --- | --- | --- |
| **Core genes** | 99% <= strains <= 100% | 2656 |
| **Soft core genes** | 95% <= strains < 99% | 316 |
| **Shell genes** | 15% <= strains < 95% | 3377 |
| **Cloud genes** | 0% <= strains < 15% | 24588 |
| **Total genes** | 0% <= strains <= 100% | 30937 |

**Supplementary Table 1.** Output from *roary* for core genome alignment of 322 samples included in the main analysis. *Roary* paramaters required genes to be present in 99% samples.

|  |  | **Died** | **Survived** | **Whole cohort** |
| --- | --- | --- | --- | --- |
| **Number of patients** |  | 38 | 275 | 313 |
| **Sex** | Female | 24 (63%) | 144 (52%) | 168 |
| **Age (years)** | Median (IQR) | 77 (67 – 88) | 74 (59 – 84) | 75 (60 – 85) |
| **Infection type** | CO-CA  CO-HA  HO-HA | 13 (34%)  14 (37%)  11 (29%) | 90 (33%)  114 (41%)  70 (25%) | 103  128  81 |
| **Source** | UTI  Non-UTI | 5 (13%)  33 (87%) | 144 (52%)  131 (48%) | 149  164 |
| **ESBL and/or AmpC** | ESBL/AmpC present | 8 (21%) | 55 (20%) | 63 |
| **Sequence type** | ST131 | 9 (24%) | 55 (20%) | 64 |

**Supplementary Table 2**. Breakdown of adult patients that died or survived at 30-days post *E. coli* bacteraemia. (Total cohort = 322; of which 318 adults). CO-CA = Community onset, community-associated; CO-HA = Community onset, healthcare-associated; HO-HA = Hospital onset, healthcare-associated.

| **Sequence type** | **count** | **%** |
| --- | --- | --- |
| 131 | 65 | 20.2 |
| 73 | 51 | 15.8 |
| 69 | 33 | 10.2 |
| 95 | 28 | 8.7 |
| 127 | 16 | 4.97 |
| 12 | 15 | 4.66 |
| 1193 | 5 | 1.55 |
| 58 | 5 | 1.55 |
| 117 | 4 | 1.24 |
| 144 | 4 | 1.24 |
| 10 | 3 | 0.932 |
| 141 | 3 | 0.932 |
| 28 | 3 | 0.932 |
| 372 | 3 | 0.932 |
| 405 | 3 | 0.932 |
| 80 | 3 | 0.932 |
| 88 | 3 | 0.932 |
| 1231 | 2 | 0.621 |
| 135 | 2 | 0.621 |
| 14 | 2 | 0.621 |
| 1406 | 2 | 0.621 |
| 162 | 2 | 0.621 |
| 362 | 2 | 0.621 |
| 367 | 2 | 0.621 |
| 38 | 2 | 0.621 |
| 404 | 2 | 0.621 |
| 428 | 2 | 0.621 |
| 453 | 2 | 0.621 |
| 533 | 2 | 0.621 |
| 648 | 2 | 0.621 |
| 681 | 2 | 0.621 |
| 83 | 2 | 0.621 |
| 1041 | 1 | 0.311 |
| 108 | 1 | 0.311 |
| 1141 | 1 | 0.311 |
| 1257 | 1 | 0.311 |
| 1266 | 1 | 0.311 |
| 1385 | 1 | 0.311 |
| 1431 | 1 | 0.311 |
| 1434 | 1 | 0.311 |
| 1485 | 1 | 0.311 |
| 1590 | 1 | 0.311 |
| 1597 | 1 | 0.311 |
| 1656 | 1 | 0.311 |
| 1876* | 1 | 0.311 |
| 191 | 1 | 0.311 |
| 2015 | 1 | 0.311 |
| 2227 | 1 | 0.311 |
| 224 | 1 | 0.311 |
| 2554 | 1 | 0.311 |
| 2599 | 1 | 0.311 |
| 3177 | 1 | 0.311 |
| 354 | 1 | 0.311 |
| 354* | 1 | 0.311 |
| 3556 | 1 | 0.311 |
| 357 | 1 | 0.311 |
| 359 | 1 | 0.311 |
| 390 | 1 | 0.311 |
| 398 | 1 | 0.311 |
| 4088 | 1 | 0.311 |
| 410 | 1 | 0.311 |
| 421 | 1 | 0.311 |
| 537 | 1 | 0.311 |
| 538 | 1 | 0.311 |
| 569 | 1 | 0.311 |
| 5891 | 1 | 0.311 |
| 59 | 1 | 0.311 |
| 607 | 1 | 0.311 |
| 625 | 1 | 0.311 |
| 640 | 1 | 0.311 |
| 652 | 1 | 0.311 |
| 6999 | 1 | 0.311 |
| 7092 | 1 | 0.311 |
| 783 | 1 | 0.311 |
| 8915 | 1 | 0.311 |
| 998 | 1 | 0.311 |
| NF* | 1 | 0.311 |

**Supplementary Table 3**. List of sequence types by frequency, as defined using *srst2*. 4 sequence types accounted for 55% of all isolates: ST131, ST73, ST69, and ST95.

| **O:H serotype** | **count** | **%** |
| --- | --- | --- |
| O25:H4 | 60 | 18.6 |
| O6:H1 | 33 | 10.2 |
| O1:H7 | 15 | 4.66 |
| O6:H31 | 15 | 4.66 |
| O17:H18 | 13 | 4.04 |
| O4:H5 | 10 | 3.11 |
| O117:H4 | 9 | 2.8 |
| O18:H7 | 8 | 2.48 |
| O2:H1 | 8 | 2.48 |
| O75:H5 | 8 | 2.48 |
| O16:H5 | 5 | 1.55 |
| O2:H4 | 5 | 1.55 |
| O4:H1 | 5 | 1.55 |
| O15:H6 | 4 | 1.24 |
| O16:H6 | 4 | 1.24 |
| O2:H6 | 4 | 1.24 |
| O2:H7 | 4 | 1.24 |
| O8:H10 | 4 | 1.24 |
| O129:H4 | 3 | 0.932 |
| O18:H1 | 3 | 0.932 |
| O75:H7 | 3 | 0.932 |
| O83:H1 | 3 | 0.932 |
| O86:H18 | 3 | 0.932 |
| O1:H6 | 2 | 0.621 |
| O102:H6 | 2 | 0.621 |
| O153:H2 | 2 | 0.621 |
| O153:H4 | 2 | 0.621 |
| O161:H4 | 2 | 0.621 |
| O18:H31 | 2 | 0.621 |
| O18:H5 | 2 | 0.621 |
| O22:H1 | 2 | 0.621 |
| O23:H16 | 2 | 0.621 |
| O51:H14 | 2 | 0.621 |
| O6:H5 | 2 | 0.621 |
| O7:H6 | 2 | 0.621 |
| O10:H12 | 1 | 0.311 |
| O102:H23 | 1 | 0.311 |
| O104:H9 | 1 | 0.311 |
| O108:H2 | 1 | 0.311 |
| O109:H10 | 1 | 0.311 |
| O11:H4 | 1 | 0.311 |
| O112:H19 | 1 | 0.311 |
| O113:H4 | 1 | 0.311 |
| O12:H4 | 1 | 0.311 |
| O120:H15 | 1 | 0.311 |
| O120:H31 | 1 | 0.311 |
| O129:H15 | 1 | 0.311 |
| O131:H12 | 1 | 0.311 |
| O134:H31 | 1 | 0.311 |
| O138:H34 | 1 | 0.311 |
| O143:H4 | 1 | 0.311 |
| O15:H1 | 1 | 0.311 |
| O15:H18 | 1 | 0.311 |
| O15:H4 | 1 | 0.311 |
| O153:H15 | 1 | 0.311 |
| O153:H25 | 1 | 0.311 |
| O153:H34 | 1 | 0.311 |
| O153:H6 | 1 | 0.311 |
| O157:H39 | 1 | 0.311 |
| O160:H21 | 1 | 0.311 |
| O160:H40 | 1 | 0.311 |
| O168:H25 | 1 | 0.311 |
| O170:H7 | 1 | 0.311 |
| O18:H14 | 1 | 0.311 |
| O2:H14 | 1 | 0.311 |
| O2:H31 | 1 | 0.311 |
| O21:H5 | 1 | 0.311 |
| O25:H1 | 1 | 0.311 |
| O25:H18 | 1 | 0.311 |
| O33:H6 | 1 | 0.311 |
| O45:H7 | 1 | 0.311 |
| O48:H20 | 1 | 0.311 |
| O5:H18 | 1 | 0.311 |
| O6:H7 | 1 | 0.311 |
| O64:H20 | 1 | 0.311 |
| O7:H15 | 1 | 0.311 |
| O7:H45 | 1 | 0.311 |
| O76:H14 | 1 | 0.311 |
| O76:H6 | 1 | 0.311 |
| O8:H19 | 1 | 0.311 |
| O8:H20 | 1 | 0.311 |
| O8:H25 | 1 | 0.311 |
| O8:H4 | 1 | 0.311 |
| O8:H8 | 1 | 0.311 |
| O83:H42 | 1 | 0.311 |
| O83:H5 | 1 | 0.311 |
| O85:H18 | 1 | 0.311 |
| O9:H19 | 1 | 0.311 |
| O9:H25 | 1 | 0.311 |
| O9:H4 | 1 | 0.311 |
| O9:H5 | 1 | 0.311 |
| O9:H9 | 1 | 0.311 |
| O96:H23 | 1 | 0.311 |
| Missing or incomplete serotype call | 11 | 3.42 |

**Supplementary Table 4**. List of O:H serotypes by frequency, as defined using *srst2*.
